# Effects of multiple-dose intranasal oxytocin treatment on social responsiveness in children with autism: A randomized, placebo-controlled trial

**DOI:** 10.1101/2022.04.20.22274106

**Authors:** Nicky Daniels, Matthijs Moerkerke, Jean Steyaert, Annelies Bamps, Edward Debbaut, Jellina Prinsen, Tiffany Tang, Stephanie Van der Donck, Bart Boets, Kaat Alaerts

## Abstract

In the past decade, intranasal administration of the neuropeptide oxytocin is increasingly explored as a new treatment for reducing the core symptoms of autism spectrum disorder (ASD). The efficacy of continual oxytocin treatment in school-aged children with ASD is, however, not well established. Using a double-blind, randomized, placebo-controlled, parallel design, the current trial explored the effects of four weeks of intranasal oxytocin treatment (12 IU, twice daily) on social functioning in pre-pubertal school-aged children (aged 8-12 years, 61 boys, 16 girls). The double-blind phase was followed by a four-week single-blind extension phase during which all participants received intranasal oxytocin. In the double-blind phase, no treatment-specific effects were identified in the primary outcome assessing social functioning (parent-rated Social Responsiveness Scale), as well as on secondary outcomes assessing repetitive behaviors, anxiety, and attachment. Exploratory moderator analyses revealed that children who received the oxytocin treatment in combination with concomitant psychosocial treatment displayed a greater benefit than those who received psychosocial treatment or oxytocin alone. A modulating effect of parents’ beliefs about allocated treatment was also identified, indicating that parents who believed their child assigned to the active treatment reported greater benefit than those who believed their child received placebo, particularly in the actual oxytocin group. Finally, participants who were allocated to receive the placebo treatment during the double-blind phase of the trial and later crossed-over to receive the active treatment during the single-blind extension phase, displayed a significant within-group improvement in social responsiveness, over and above the placebo-induced improvements noted in the first phase. While no overall treatment-specific improvements were identified, our results provide important indications that clinical efficacy can be augmented when oxytocin administration is paired with targeted psychosocial interventions that similarly stimulate socio-communicative behaviors. Future trials are urged to further elucidate the potential of embedding oxytocin treatment within a socially stimulating context.

## Introduction

Autism spectrum disorder (ASD) is a neurodevelopmental condition characterized by impairments in social communication and interaction, combined with restricted and repetitive behaviors and interests (American Psychiatric Association, 2013). Thus far, the means of treatment of ASD’s core symptoms are primarily based on behavioral interventions (e.g., stimulation of social communication, lessening the impairment due to restricted and repetitive behaviors), since biomedical or pharmacological therapies targeting social impairment or repetitive behaviors are largely unproven.

In the past decade, intranasal administration of the neuropeptide oxytocin (OT) has been increasingly explored as a new treatment option for reducing ASD’s social symptoms (recently summarized in Huang et al., 2021). OT is an endogenous neuropeptide that is mainly produced in paraventricular nuclei of the hypothalamus. In the brain, OT acts as an important neuromodulator for a broad range of affiliative and prosocial behaviors, including interpersonal bonding, social attunement and attachment (Bakermans-Kranenburg & van IJzendoorn, 2013; Bartz et al., 2011; Jurek & Neumann, 2018), presumably mediated through its postulated top-down enhancing effect on ‘social salience’ and bottom-up effect on regulating (social) stress and anxiety (Guastella & Hickie, 2016; Shamay-Tsoory & Abu-Akel, 2016).

Following a myriad of single-dose proof-of-principle studies (Alvares et al., 2017; Huang et al., 2021), an initial multiple-dose pilot study assessed the safety and efficacy of six weeks of chronic intranasal OT treatment on core autism symptoms in 19 adults with ASD (10 receiving OT, 9 receiving placebo), and showed improved emotion recognition and quality of life, and tentative improvements in repetitive behaviors after OT treatment (Anagnostou et al., 2012). Later, Kosaka et al. (2016) showed significant improvements on the Clinical Global Impression-Improvement scale after twelve weeks of OT treatment in adult men with ASD, albeit only in the subgroup of participants receiving the high-dose treatment (32 IU/day; n = 13), and not in the low-dose (16 IU; n = 15) or placebo groups (n = 16). In an exploratory cross-over study by Watanabe et al. (2015), the effects of six weeks of daily intranasal OT administration on core autism characteristics were studied in 20 adult men with ASD, and significant improvements in social reciprocity and social functioning (social-judgement task) were identified. Yamasue et al. (2020) conducted a confirmatory trial with an identical protocol as Watanabe et al. (2015) in 106 adult men with ASD (53 OT / 53 placebo). While these authors identified significant improvements in terms of repetitive behaviors, the effects on social reciprocity and social functioning could not be replicated. Bernaerts et al. (2020) extended these observations in an exploratory sample of 40 young adult men with ASD (22 OT / 18 placebo), demonstrating long-term improvements in repetitive behaviors and feelings of attachment after a four-week course which outlasted the period of administration till one year post-treatment.

Given that ASD is an early-onset neurodevelopmental condition, it is important to extend these insights to pediatric populations, allowing evaluations of OT treatment efficacy within an early developmental window and whether it can be facilitatory for enriching social behaviors and experiences from an early age onwards. To date, a handful of trials explored the effects of multiple-dose OT administration in children with ASD. Two initial trials reported a consistent pattern of results, indicating improvements in the social domain (parent-reported social responsiveness) after five weeks of intranasal OT treatment in 3-to-6-year-old children (n = 31; cross-over; Yatawara et al., 2016) and after four weeks of treatment in 6-to-12-year-old children with ASD (14 OT / 18 placebo; Parker et al., 2017). No significant improvements on core autism symptoms were demonstrated, however, after an eight-week OT treatment in adolescent boys with ASD (26 OT / 24 placebo; 12-18 years; Guastella et al., 2015) or in a preliminary 12-week administration trial encompassing a broad age range of 5-to-17-year-old children (8 OT / 10 placebo) with Phelan-McDermid syndrome (characterized by ASD symptoms; Fastman et al., 2021). Also, in a recent confirmatory trial including 3-to-17-year-old children with ASD (139 OT / 138 placebo) and an age-adjusted dosing scheme ranging from 8-80 IU, no improvements on outcomes of social functioning were evident after 24 weeks of OT treatment (Sikich et al., 2021).

Several factors have been put forward to understand these inconsistent results, ranging from heterogeneity in trial design (e.g., parallel versus cross-over design, adopted outcomes, dosing schema) to variation in participant characteristics. For instance, the well-powered confirmatory trial by Sikich et al. (2021) covered a broad age range (3-17 years), encompassing a critical period of pubertal development, which could have rendered heterogeneity due to differential physiologic effects of OT during different developmental stages (Geschwind, 2021).

Here, results are presented from a single-center, randomized, double-blind, placebo-controlled clinical trial (RCT with parallel design), testing efficacy on improvement in social functioning and safety of multiple-dose OT treatment (four weeks of twice daily intranasal administration of 12 IU) in a representative sample of 8-to-12-year-old children with ASD (40 OT / 40 placebo). Accordingly, this is the largest trial to date, examining OT treatment effects in a relatively strict age range of pre-pubertal, school-aged children, aged 8 to 12 years, thereby allowing to overcome some of the raised issues regarding sample heterogeneity. Further, following prior observations of long-lasting retention effects of OT treatment in adults with ASD (Bernaerts et al., 2020), the current trial also included a follow-up session four weeks after cessation of the daily OT administrations, testing the possibility of crucial retention effects in the current pediatric sample.

## Methods

### 2.1. General study design

A single-center, two-arm, double-blind, randomized, placebo-controlled parallel study was performed at the Leuven University Hospital (Leuven, Belgium) to assess effects of four weeks of twice daily intranasal administration of OT on core autism characteristics, social anxiety, and experience of attachment in school-aged children with ASD. The double-blind phase (phase I) was followed by a four-week single-blind extension phase (phase II) during which all participants received intranasal OT. In both phases, treatment effects were assessed immediately after the four-week treatment (post) and at a follow-up session, four weeks after cessation of the daily administrations (follow-up). See **Figure 1**, CONSORT Flow diagram for number of participants randomized and analyzed.

**Figure 1.**
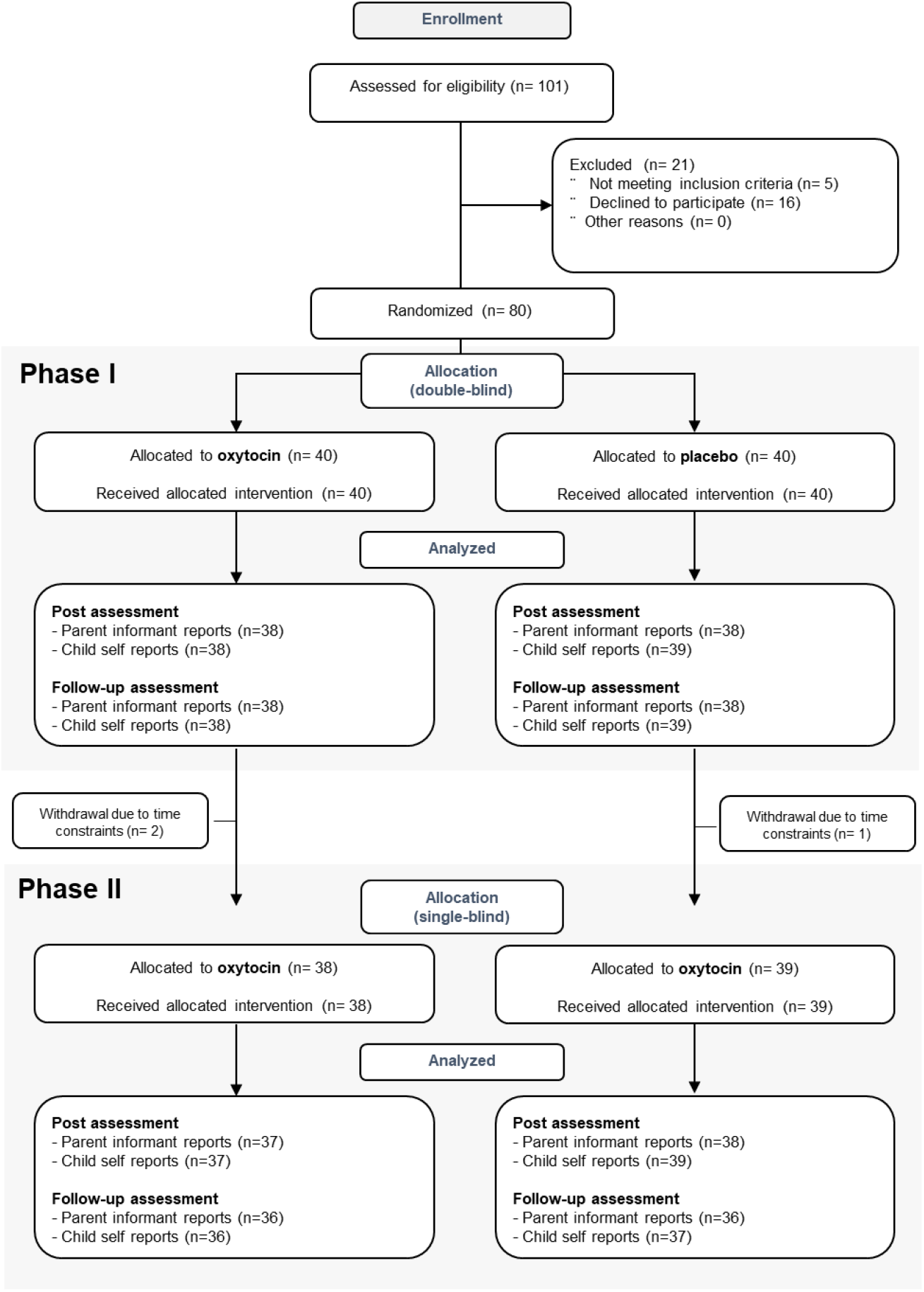
CONSORT flow diagram of participants in the trial. Participants first underwent a double-blind phase (phase I) during which they were allocated to receive either oxytocin or placebo (four weeks of twice daily intranasal administration), followed by a four-week single-blind extension phase (phase II) during which all participants received four weeks of intranasal oxytocin. In both phases, treatment effects were assessed immediately after the four-week treatment (post) and at a follow-up session four weeks after cessation of the daily administrations (follow-up). For each assessment session, completed assessments are indicated separately for parent informant- and child self-reports.

Written informed consent from the parents and assent from the child were obtained prior to the study. Consent forms and study design were approved by the local Ethics Committee for Biomedical Research at the University of Leuven, KU Leuven (S61358) in accordance with The Code of Ethics of the World Medical Association (Declaration of Helsinki). The trial was registered at the European Clinical Trial Registry (Eudract 2018-000769-35) and the Belgian Federal Agency for Medicines and Health products.

### 2.2. Participants

Children with a formal diagnosis of ASD were recruited through the Autism Expertise Centre at the Leuven University Hospital between July 2019 and January 2021. The diagnosis was established by a multidisciplinary neuropediatric team based on the strict criteria of the DSM-5 (Diagnostic and Statistical Manual of Mental Disorders; American Psychiatric Association, 2013). Prior to randomization, the Autism Diagnostic Observation Schedule (ADOS-2; Lord et al., 2012) and estimates of intelligence (four subtests of the Wechsler Intelligence Scale for Children, Fifth Edition, Dutch version; Wechsler, 2018) were acquired (**Table 1**). Principal inclusion criteria comprised a clinical diagnosis of ASD, age (8-12 years old), intelligence quotient (IQ) above 70, native Dutch speaker, a stable background treatment for at least four weeks prior to the screening and no anticipated changes during the trial. Only premenstrual girls were included. Principal criteria for exclusion comprised any neurological (e.g., stroke, epilepsy, concussion) or significant physical disorder (liver, renal, cardiac pathology) or prior treatment with OT (**Supplementary Table 1**). The presence of comorbid psychiatric disorders, current psychoactive medication use, and concomitant participation in psychosocial therapies were screened for and logged (see **Table 1 and Supplementary Table 2**).

**Table 1.**
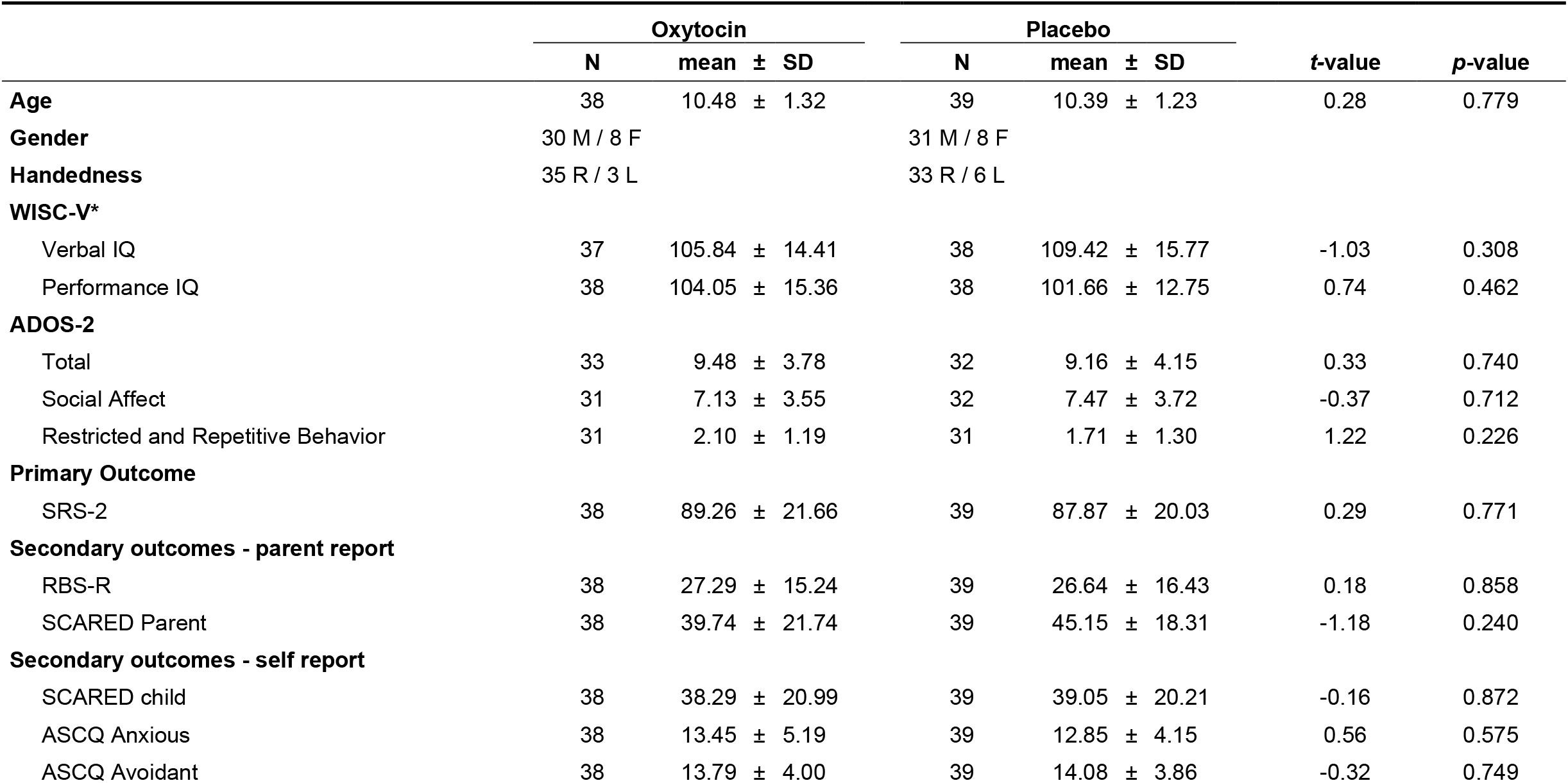
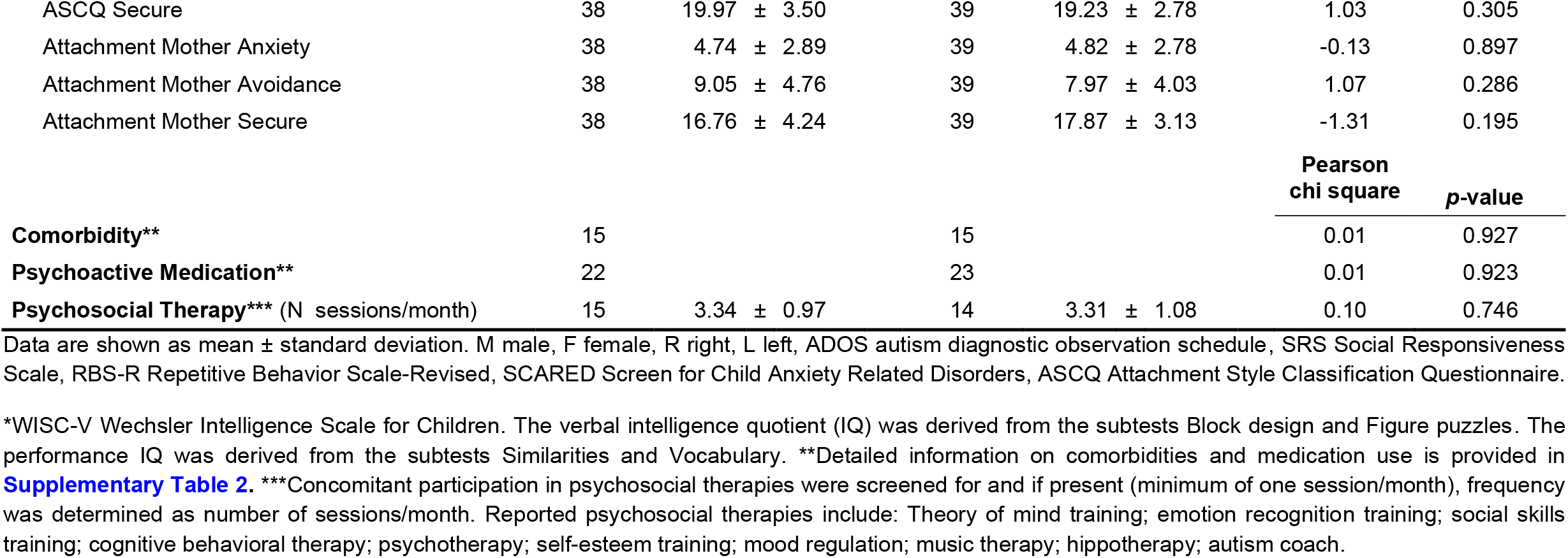
Demographic characteristics of the trial participants at baseline. Mean baseline scores listed separately for the oxytocin and placebo treatment groups. *T*- and *p*-values correspond to independent sample *t*-tests assessing between-group differences in baseline scores.

|  | Oxytocin |  |  | Placebo |  |  | *t*-value | *p*-value |
| --- | --- | --- | --- | --- | --- | --- | --- | --- |
|  | N | mean | ± SD | N | mean | ± SD |  |  |
| **Age** | 38 | 10.48 | ± 1.32 | 39 | 10.39 | ± 1.23 | 0.28 | 0.779 |
| **Gender** | 30 M / 8 F |  |  | 31 M / 8 F |  |  |  |  |
| **Handedness** | 35 R / 3 L |  |  | 33 R / 6 L |  |  |  |  |
| **WISC-V\*** |  |  |  |  |  |  |  |  |
| Verbal IQ | 37 | 105.84 | ± 14.41 | 38 | 109.42 | ± 15.77 | -1.03 | 0.308 |
| Performance IQ | 38 | 104.05 | ± 15.36 | 38 | 101.66 | ± 12.75 | 0.74 | 0.462 |
| **ADOS-2** |  |  |  |  |  |  |  |  |
| Total | 33 | 9.48 | ± 3.78 | 32 | 9.16 | ± 4.15 | 0.33 | 0.740 |
| Social Affect | 31 | 7.13 | ± 3.55 | 32 | 7.47 | ± 3.72 | -0.37 | 0.712 |
| Restricted and Repetitive Behavior | 31 | 2.10 | ± 1.19 | 31 | 1.71 | ± 1.30 | 1.22 | 0.226 |
| **Primary Outcome** |  |  |  |  |  |  |  |  |
| SRS-2 | 38 | 89.26 | ± 21.66 | 39 | 87.87 | ± 20.03 | 0.29 | 0.771 |
| **Secondary outcomes - parent report** |  |  |  |  |  |  |  |  |
| RBS-R | 38 | 27.29 | ± 15.24 | 39 | 26.64 | ± 16.43 | 0.18 | 0.858 |
| SCARED Parent | 38 | 39.74 | ± 21.74 | 39 | 45.15 | ± 18.31 | -1.18 | 0.240 |
| **Secondary outcomes - self report** |  |  |  |  |  |  |  |  |
| SCARED child | 38 | 38.29 | ± 20.99 | 39 | 39.05 | ± 20.21 | -0.16 | 0.872 |
| ASCQ Anxious | 38 | 13.45 | ± 5.19 | 39 | 12.85 | ± 4.15 | 0.56 | 0.575 |
| ASCQ Avoidant | 38 | 13.79 | ± 4.00 | 39 | 14.08 | ± 3.86 | -0.32 | 0.749 |

|  |  |  |  |  |  |  |
| --- | --- | --- | --- | --- | --- | --- |
| ASCQ Secure | 38 | 19.97 ± 3.50 | 39 | 19.23 ± 2.78 | 1.03 | 0.305 |
| Attachment Mother Anxiety | 38 | 4.74 ± 2.89 | 39 | 4.82 ± 2.78 | -0.13 | 0.897 |
| Attachment Mother Avoidance | 38 | 9.05 ± 4.76 | 39 | 7.97 ± 4.03 | 1.07 | 0.286 |
| Attachment Mother Secure | 38 | 16.76 ± 4.24 | 39 | 17.87 ± 3.13 | -1.31 | 0.195 |

|  |  |  |  |  | Pearson<br>chi square | p-value |
| --- | --- | --- | --- | --- | --- | --- |
| **Comorbidity\*\*** | 15 |  | 15 |  | 0.01 | 0.927 |
| **Psychoactive Medication\*\*** | 22 |  | 23 |  | 0.01 | 0.923 |
| **Psychosocial Therapy\*\*\*** (N sessions/month) | 15 | 3.34 ± 0.97 | 14 | 3.31 ± 1.08 | 0.10 | 0.746 |

Data are shown as mean ± standard deviation. M male, F female, R right, L left, ADOS autism diagnostic observation schedule, SRS Social Responsiveness Scale, RBS-R Repetitive Behavior Scale-Revised, SCARED Screen for Child Anxiety Related Disorders, ASCQ Attachment Style Classification Questionnaire.
\*WISC-V Wechsler Intelligence Scale for Children. The verbal intelligence quotient (IQ) was derived from the subtests Block design and Figure puzzles. The performance IQ was derived from the subtests Similarities and Vocabulary. \*\*Detailed information on comorbidities and medication use is provided in Supplementary Table 2. \*\*\*Concomitant participation in psychosocial therapies were screened for and if present (minimum of one session/month), frequency was determined as number of sessions/month. Reported psychosocial therapies include: Theory of mind training; emotion recognition training; social skills training; cognitive behavioral therapy; psychotherapy; self-esteem training; mood regulation; music therapy; hippotherapy; autism coach.

A sample size of 40 participants in each treatment group was determined to be able to detect a medium effect size (*d* = 0.60) with α = 0.05 and 80% power, corresponding to effect sizes previously reported in the four-week oxytocin trial with school-aged children by Parker et al., 2017.

### 2.3. Intervention

#### Study medication

Participants were randomized to receive OT (Syntocinon®, Sigma-tau) or placebo nasal sprays, administered in identical blinded amber 10 ml glass bottles with metered pump. The placebo spray consisted of all the ingredients used in the active solution except the OT compound. Nasal spray preparation, packaging, blinding and randomization (permuted-block randomization, RITA software; Pahlke et al., 2004) was performed by the pharmacy of Heidelberg University Hospital (Germany). Participants were randomly assigned in a 1:1 ratio, with stratification according to gender. During the initial double-blind phase (phase I), all research staff conducting the trial, participants and their parents were blinded to treatment allocation. During the subsequent single-blind extension phase (phase II), experimenters were aware that all participants received intranasal OT, but participants and parents were still fully blinded regarding treatment allocation.

#### Dosing

Children (assisted by their parents) were asked to self-administer a daily dose of 2 × 12 IU nasal spray or placebo equivalent (3 puffs of 2 IU in each nostril), 12 IU in the morning and 12 IU in the afternoon, during 28 consecutive days during the initial double-blind phase (phase I), and for another 28 days during the single-blind extension phase (phase II). Participants received clear instructions about use of the nasal sprays (based on Guastella et al., 2013) through a demonstration together with the experimenter.

#### Compliance monitoring

Compliance was assured using a daily medication diary that recorded date and time of administration (phase I percentage compliance; OT: 96.75 ± 5.26%; placebo: 96.11 ± 5.29 %; *t*(74) = .52, *p* = .603; phase II percentage compliance; OT-first: 94.55 ± 11.69%; placebo-first: 92.98 ± 13.92 %; *t*(74) = .53, *p* = .597). The total amount of administered fluid was also monitored (phase I: OT: 14.86 ± 2.37 ml; PL: 13.79 ± 2.35 ml; *t*(75) = 2.00, *p* = .050; phase II: OT-first: 13.72 ± 3.47 ml; placebo-first: 12.83 ± 3.52 ml; *t*(74) = 1.10, *p* = .275).

#### Side effects

During the course of the treatment, participants were screened for potential adverse events (weekly parent report) or changes in affect and arousal (daily diary by child and parent). Overall, reports of side effects were minimal and not treatment-specific (see **Supplementary Tables 3 and 4**).

#### Parent reported treatment beliefs

At the end of each trial phase (I and II), parents reported beliefs about treatment allocation (see **Results** and **Figure 3**). In the double-blind phase (phase I), the proportion of parents that believed their child had received the OT treatment was similar in both treatment arms: 39.5% in the OT group, 35.9% in the placebo group (*p* = .75). In the OT group 18.4% of parents indicated to ‘have no explicit belief’ about treatment allocation versus 10.3% in the placebo group. In the single blind phase (phase II), during which all participants received the actual OT treatment, 51.9% of the parents believed their child received the OT treatment, 35.1% believed their child received the placebo treatment and 13.0% indicated to ‘have no explicit belief’.

### 2.4. Outcome Measures

The primary outcome measure was change from baseline in parent-rated social responsiveness on the Social Responsiveness Scale-Children, second edition (SRS-2; Constantino & Gruber, 2012; Roeyers et al., 2015). which comprises four subscales examining social communication, social awareness, social motivation, and rigidity/repetitiveness, using a four-point Likert-scale (65 items). Higher scores indicate greater deficit.

Secondary outcome measures included changes from baseline in parent-rated repetitive behaviors (Repetitive Behavior Scale-Revised (RBS-R; Bodfish et al., 2000), self and parent-rated presence of anxiety symptoms (Screen for Child Anxiety Related Emotional Disorders (SCARED-NL: Muris et al., 2007), and changes from baseline in constructs of self-rated attachment towards their mother (Attachment Questionnaire child-report; Bosmans et al., 2014) and peers (Attachment Style Classification Questionnaire child-report; Finzi et al., 2000) (see also **Supplementary Table 5**).

All outcomes were assessed five times: (i) at baseline, (ii) immediately after the four-week double-blind treatment (phase I - post); (iii) at a follow-up session, four weeks after cessation of the double-blind treatment (phase I - follow-up); (iv) immediately after the four-week single-blind treatment (phase II - post); and (v) at a follow-up session four weeks after cessation of the single-blind treatment (phase II - follow-up). Post sessions were scheduled approximately 24h after the last administration, follow-up sessions within 28 ± 7 days.

### 2.5 Data Analysis

Analyses were performed using a modified intention-to-treat approach that included all randomized participants who completed the baseline session and at least one post or follow-up session (see **Figure 1, CONSORT flow-chart**). All statistics were executed with Statistica 14 (Tibco Software Inc.).

First, as outlined in **Table 1**, possible baseline differences between treatment groups on the primary or secondary outcomes were assessed using independent sample t-tests, but no statistically significant differences were identified. Next, between-group differences in treatment responses of *phase I (double-blind)* on the primary and secondary outcome measures were assessed, by subjecting change from baseline scores of the post and follow-up session to independent sample t-tests. Additionally, single-sample *t*-tests were adopted to assess within-group changes (compared to baseline) in the OT and placebo group separately.

Subsequent exploratory analyses of the primary outcome were performed to investigate the potential influence of possible moderator variables on phase I treatment outcome. To do so, change from baseline scores of the primary outcome (SRS-2) were subjected to general linear models with the within-subject factor ‘assessment session’ (post, follow-up) and the between-subject factors ‘treatment’ (OT, placebo) and specific moderator variables. Separate models were constructed to assess the modulating effect of concomitant psychosocial treatments (present, not present); medication use (present, not present; as listed in **Table 1**); and parent reported beliefs (OT, placebo).

To evaluate the effect of phase, within-subject changes from *phase I (double-blind) to phase II (single-blind extension)* were assessed, across groups and separately within the OT-first and placebo-first groups. To do so, change from baseline scores of the primary outcome (SRS-2) were subjected to general linear models with the within-subject factors ‘assessment session’ (post, follow-up) and ‘phase’ (phase I, phase II).

Finally, to assess whether the magnitude of the observed change from baseline scores at the last session of the trial were reliable for individual participants (more than can be expected by measurement error), the Reliable Change Index (RCI) as proposed by Jacobson et al. (1999) was calculated, based on the test-retest reliability of the adopted Dutch parent-reported SRS scale (Cronbach’s alpha = .94; Roeyers et al., 2015). Change scores higher than the RCI-value (14.8) were considered reliable.

## Results

### Double-blind phase (phase I)

Between-group analyses revealed no significant effect of treatment on the primary outcome parent-reported social responsiveness (SRS-2), either immediately post-treatment (*p* = .839), or at the follow-up session, four weeks after cessation of the daily nasal spray administrations (*p* = .626) (see **Table 2** and **Supplementary Table 6** for the raw scores). Both groups displayed similar significant pre-to-post-treatment improvements in social responsiveness (reduced SRS-2 scores) immediately after treatment (OT: *p* = .017; placebo: *p* = .009) and at the follow-up session (OT: *p* = .001; placebo: *p* = .017). A similar pattern of non-treatment-specific improvements was evident for the secondary outcomes (**Table 2**).

**Table 2.**
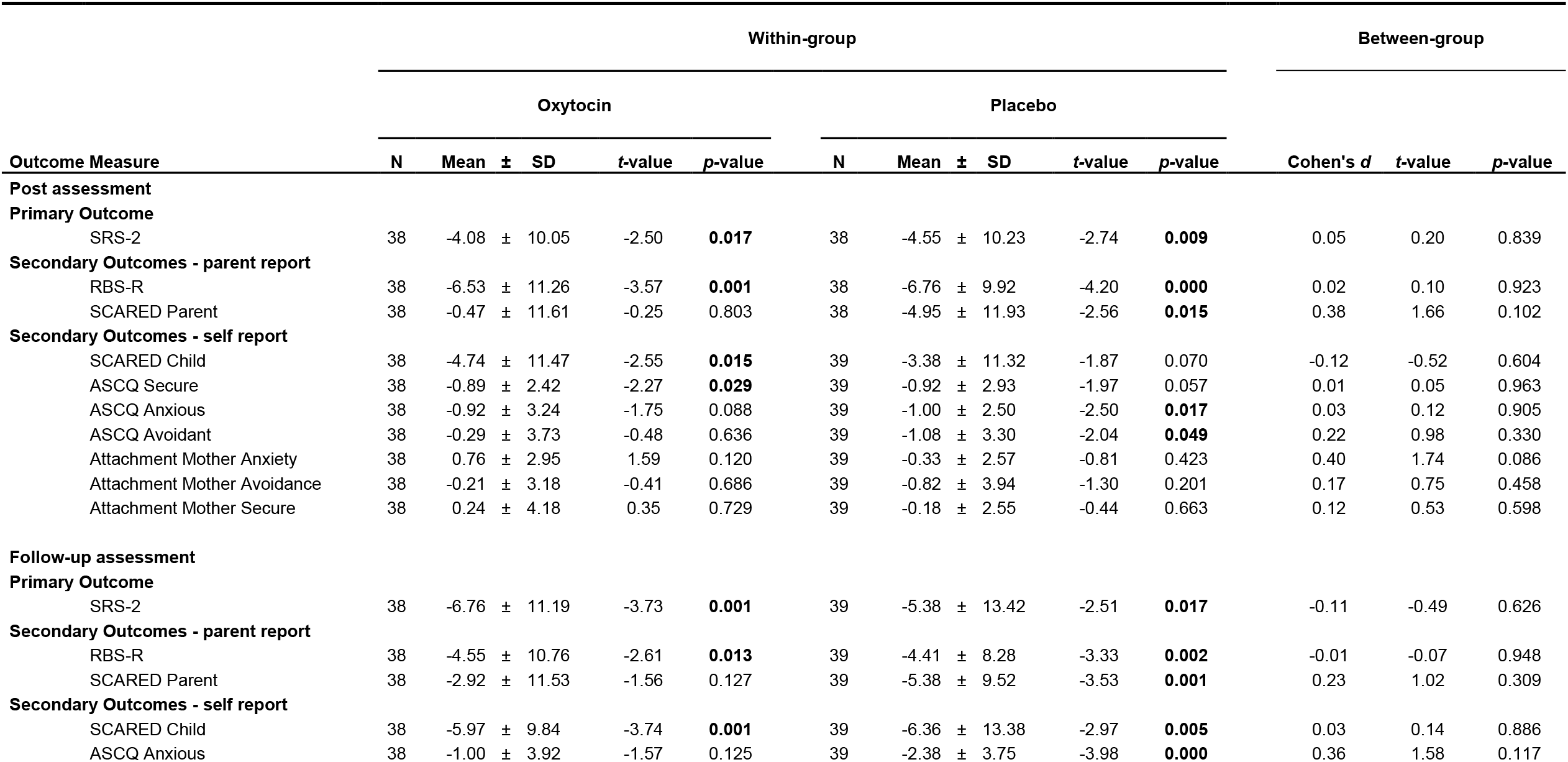
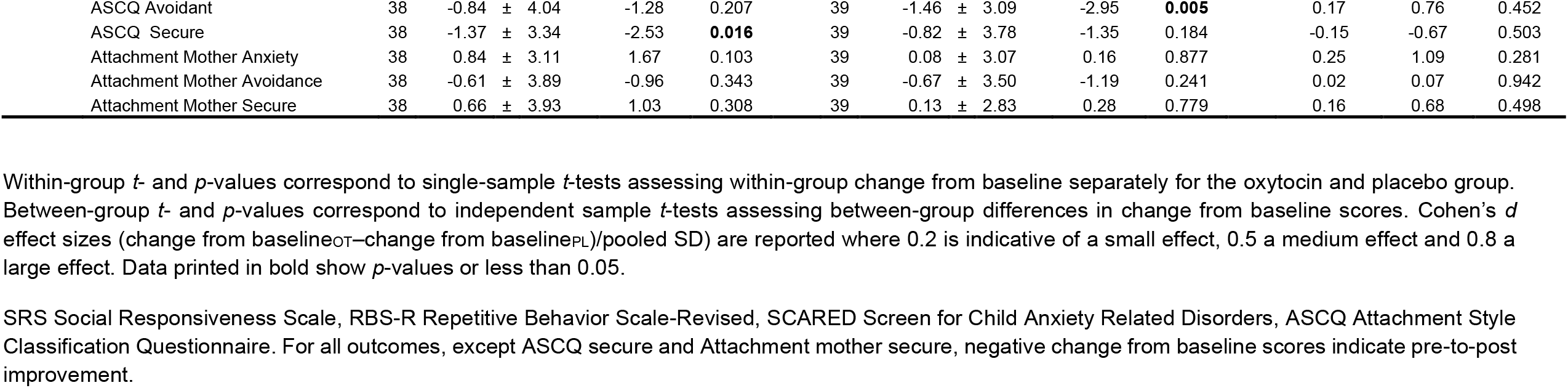
Effects of oxytocin treatment on primary and secondary outcome measures of the double-blind phase I. Mean change from baseline scores are listed separately for the oxytocin and placebo treatment groups, and separately for the post assessment session (immediately after the four-week treatment) and the follow-up assessment (four weeks after cessation of the treatment).

Interestingly, exploratory moderator analyses showed a significant interaction between treatment and the presence of **concomitant psychosocial treatment** (*F*(1,72) = 6.87; *p* = .011; **Figure 2**), indicating that, across assessment sessions (post, follow-up), participants who received the OT treatment combined with psychosocial treatment displayed greater benefits compared to children receiving only psychosocial treatment (combined with placebo) (*t*(26) = 2.40; *p* = .012, one-tailed) or only the OT treatment (*t*(36) = 1.90; *p* = .033, one-tailed). In children without psychosocial treatment, OT treatment responses were not significantly different from those in the placebo group (*t*(46) = 1.40; *p* = .084, one-tailed). None of the other main or interaction effects (e.g., with assessment session) were significant (all, *p* > .05).

**Figure 2.**
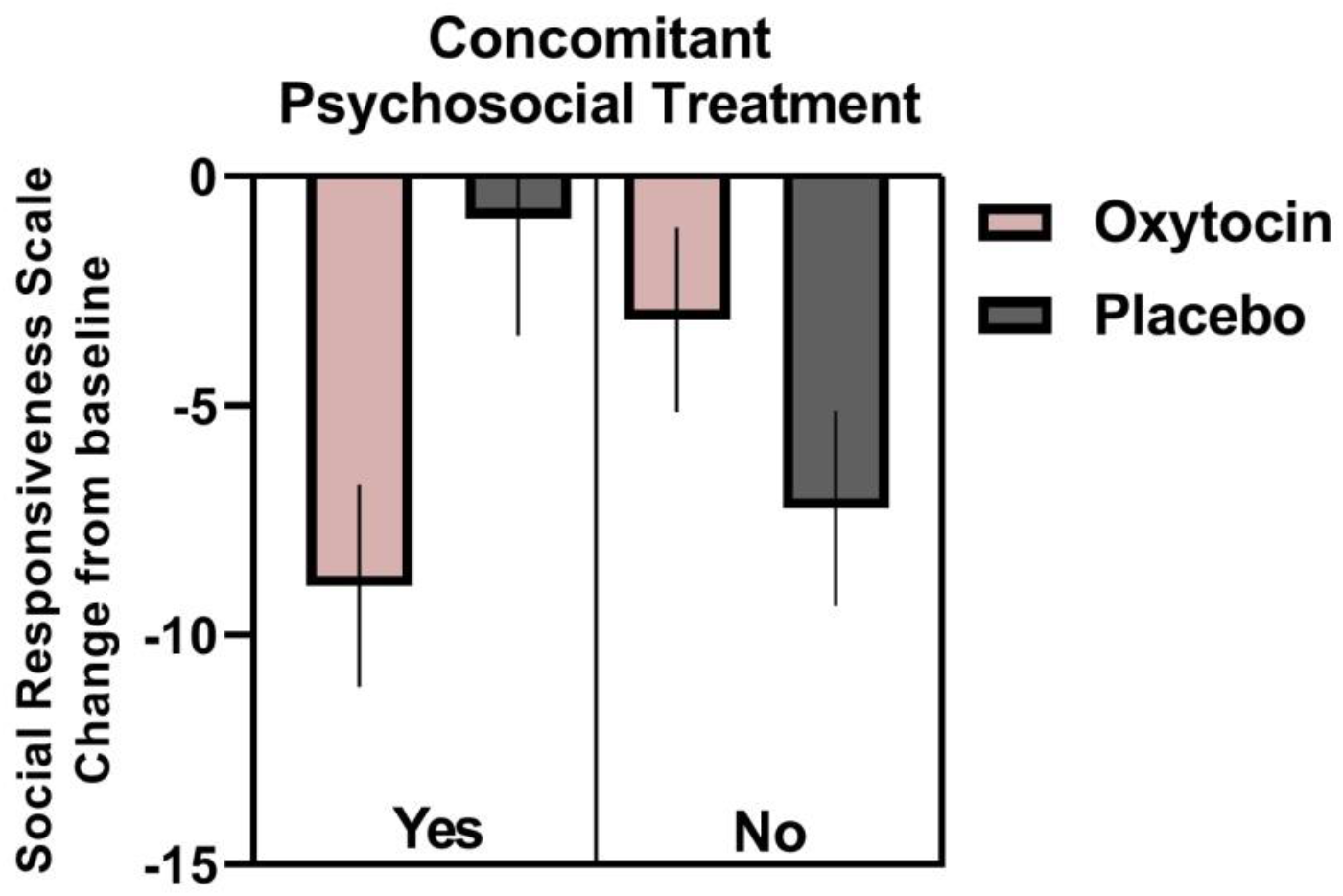
Change in treatment responses according to the presence of concomitant psychosocial treatment. Visualization of changes from baseline in parent-reported social responsiveness (SRS) of the double-blind phase (phase I), separately for children receiving only the oxytocin (n = 23) or placebo (n = 25) treatment and children receiving oxytocin (n = 15) or placebo (n = 13) treatment in combination with concomitant psychosocial treatment (pooled across the immediate post and four-week follow-up session). Lower scores indicate improvement. Vertical bars denote ± standard errors.

In terms of modulating effects of **parent reported beliefs**, a trend-level interaction with ‘assessment session’ was evident (*F*(1,61) = 3.22; *p* = .077), indicating that the parents’ own belief about allocated treatment differentially modulated treatment responses at the post and follow-up assessment session (**Figure 3**). Direct exploration of this effect, separately for each treatment group, showed that for participants receiving the actual OT treatment, parents’ own belief moderated treatment immediately post-treatment (*t*(29) = -3.18; *p* = .001, one-tailed), but no longer at the follow-up session (*t*(29) = -1.02; *p* = .158, one-tailed). Specifically, parents who believed their child had received OT, reported significantly greater improvements in social responsiveness immediately post-treatment, compared to those who believed their child had received placebo. In the placebo group, no significant modulations of treatment responses were evident either immediately post-treatment (*t*(32) = -.32; *p* = .374, one-tailed) or at the follow-up session (*t*(32) = .37; *p* =.358, one-tailed). None of the other main or interaction effects were significant (all, *p* > .05). Further, for **concomitant medication use**, no significant modulating effects were evident (all, *p* > .148).

**Figure 3.**
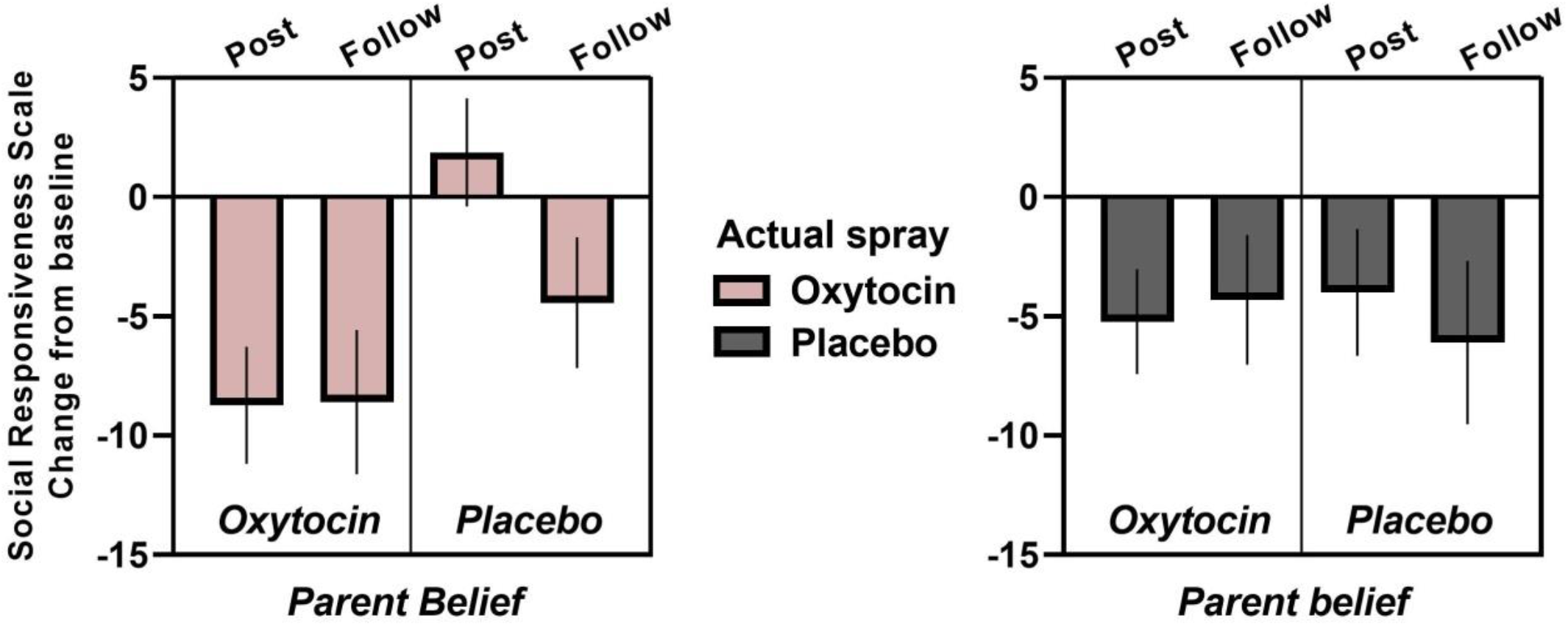
Change in treatment responses according to parent reported beliefs about the allocated treatment. Visualization of changes from baseline in parent-reported social responsiveness (SRS) of the double-blind phase (phase I) at the post (immediately after the four week treatment) and four-week follow-up session, separately for each treatment group (actual spray: oxytocin or placebo) and according to parent reported beliefs about the allocated treatment (oxytocin or placebo): oxytocin_spray_/oxytocin_belief_: n = 15; oxytocin_spray_/placebo_belief_: n = 16; placebo_spray_/oxytocin_belief_: n = 13; placebo_spray_/placebo_belief_: n = 21. Lower scores indicate improvement. Vertical bars denote ± standard errors.

### Single-blind extension phase (phase II)

Examination of within-subject changes from phase I to phase II yielded a significant effect of ‘phase’ (*F*(1,70) = 12.94; *p* < .001), but no ‘phase x treatment interaction’ (*F*(1,70) = 2.24; *p* = .14), indicating a further improvement in social responsiveness across treatment groups from phase I to phase II, (**Figure 4** and **Supplementary Table 6** for the raw scores). The main effects of treatment and assessment session were not significant (*p* > .05).

**Figure 4.**
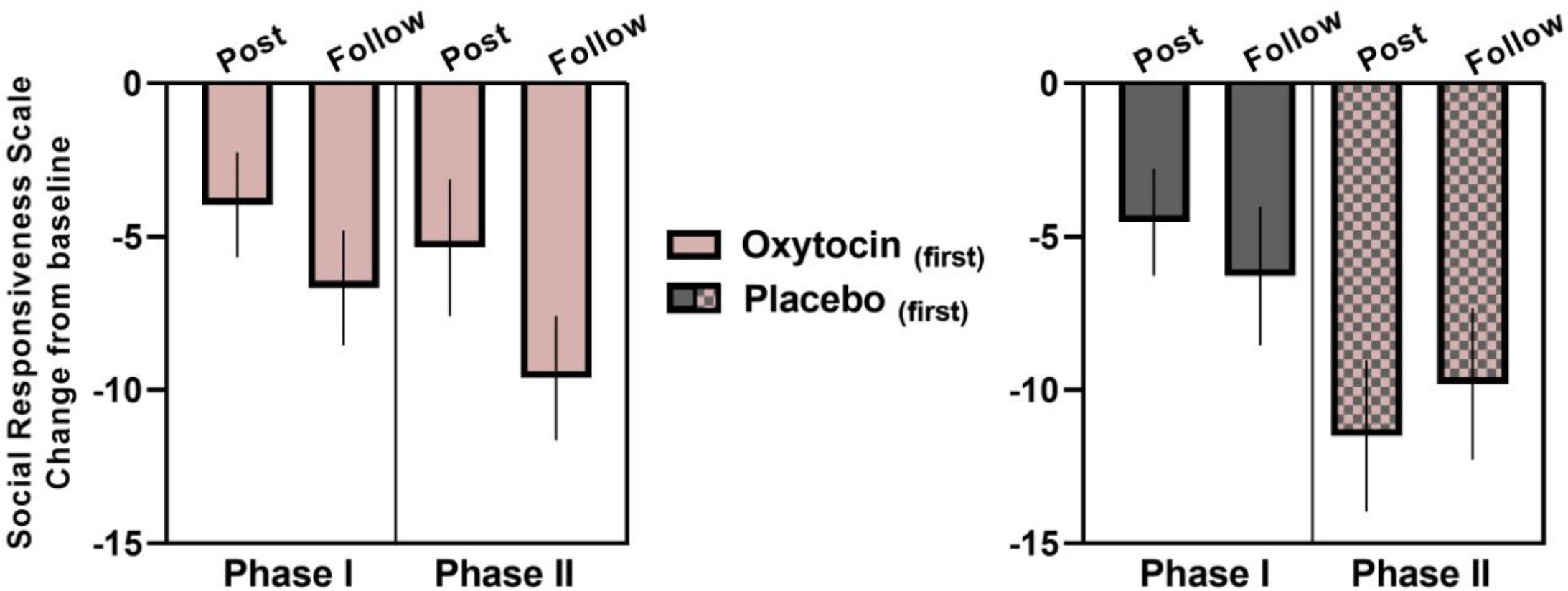
Treatment responses of the single-blind extension phase (phase II). Visualization of changes from baseline in caregiver-reported social responsiveness (SRS) of the double-blind phase (phase I) and the single-blind extension phase (phase II), separately for each original treatment group (oxytocin-first, placebo-first) and assessment session (immediate post and four-week follow-up). Lower scores indicate improvement. Vertical bars denote ± standard errors.

When examined separately for each treatment group, the effect of phase was particularly strong in the placebo-first group (*F*(1,35) = 14.54; *p* < .001), indicating that for children who crossed over from placebo (in phase I) to OT treatment (in phase II), improvements in social responsiveness were significantly more pronounced in phase II, during which the child received the actual OT treatment (**Figure 4, right panel**).

Within the OT-first group, only non-significant within-subject changes from phase I to phase II were noted (*F*(1,35) = 1.99; *p* = .17), indicating that OT treatment effects of phase I were not significantly augmented by receiving the additional four-week OT treatment of phase II. Analysis of the OT-first group did reveal a significant effect of ‘session’, indicating that irrespective of phase, treatment-related improvements were more pronounced at the follow-up session, compared to the post session (*F*(1,35) = 6.11; *p* = .018), with maximal treatment responses at the last assessment session of the trial (four-week follow-up of phase II; **Figure 4, left panel**).

Accordingly, at the last session of the trial, both the OT-first (receiving a total of eight weeks of OT treatment) and the placebo-first group (receiving a total of four weeks of OT treatment) displayed significant pre-to-post improvements in social responsiveness (OT-first; pre-post change: -9.61 ± 12.18; *t*(35)= -4.74; *p* < .001; (placebo-first; pre-post change: -9.81 ± 14.83; *t*(35)= -3.97; *p* < .001). In the OT-first group, 27 (out of 36: 75%) participants displayed a pre-to-post improvement, and this change was identified to be reliable for 12 participants (higher than the Reliable Change Index: >14.8). Similarly, also in the PL-first group, 27 (out of 36: 75%) participants displayed a pre-to-post improvement at the last session of the trial, which was reliable for 11 participants.

## Discussion

The current pediatric trial with pre-pubertal school-aged children with ASD demonstrated no significant treatment-specific effects of four weeks of intranasal OT administration on the primary outcome, assessing parent-rated social responsiveness (SRS-2), nor on the secondary outcomes assessing parent and child self-reports of repetitive behaviors, anxiety, and attachment. Both the OT and the placebo group displayed similar improvements, both immediately after the multiple-dose treatment and at the four-week follow-up session. Notably, exploratory analyses showed that children who received the OT treatment in combination with concomitant psychosocial treatment displayed a greater improvement in social responsiveness than those who received psychosocial treatment or OT alone. A modulating effect of parents’ belief about allocated treatment was also identified, indicating that parents who believed their child had been assigned the active treatment reported greater benefit than those who believed their child received placebo, particularly in the experimental group receiving actual OT. Finally, participants who were allocated to receive the placebo treatment during the first double-blind phase of the trial and later crossed-over to receive the active treatment during the second (single-blind) phase, displayed a significant improvement in social responsiveness, over and above the ‘placebo-induced’ improvement noted in the first phase. The next sections will address these observations in more detail.

The results of earlier continual OT trials in children with ASD have been equivocal: some with beneficial outcomes (Parker et al., 2017; Yatawara et al., 2016), while others without significant effect (Fastman et al., 2021; Guastella et al., 2015; Sikich et al., 2021). While it is difficult to pinpoint the different factors contributing to variability in study results, several key differences in adopted dosing schema, trial design, and participant demographics have been put forward as important moderators. Furthermore, the particular context in which the OT treatment is administered is also increasingly put forward as a vital factor for understanding variability in treatment responses within and across studies. Initial single-dose administration studies already noted that acute effects of OT can be modulated by contextual factors, indicating for instance that OT-induced facilitation of cooperation and trust is most pronounced towards in-group members, but is diminished, absent or even reversed towards out-group members, particularly within threat-emanating contexts (de Dreu et al., 2010; Mikolajczak et al., 2010). Also, in an early study by Heinrichs et al (2003), stress-reducing effects of OT were significantly augmented when accompanied by a supportive context (i.e., social support from a friend).

Against this background, it has been theorized that OT may indeed open a ‘window of opportunity’ to enhance prosocial behavior, but its potential can only be fully realized when OT treatment is paired with a supportive context, such as effective concomitant behavioral interventions that can support social skill development and improve prosocial behavior (Ford & Young, 2021; Geschwind, 2021). In line with this notion, exploratory assessments within our current trial revealed a significant synergetic modulation of treatment response related to the presence of **concomitant psychosocial treatment** during the course of the OT trial (screened through parent report, see **Table 1**), indicating maximal treatment effects in children receiving the OT treatment in combination with ongoing psychosocial treatment. Administration of OT as an adjunct to other therapeutic approaches has been explored before. For example, in a study with schizophrenic patients, a six-week (12 session) social cognition training was combined with OT administration (shortly before the start of each session), and a significant improvement in empathic accuracy was observed (Davis et al., 2014). Also, in patients with social anxiety disorders, OT treatment administered as an adjunct to 4 sessions of public speaking-exposure therapy induced significant improvements in mental representations of the self (Guastella et al., 2009). While preliminary, a pilot 6-week OT administration study in which parents were stimulated to systematically engage with their child in a positive social interaction or play session in the first hour after spray administration, yielded unanimously positive treatment outcomes in 3-to-8-year-old children with ASD (n = 46), both in terms of social improvements and repetitive behaviors (Le et al., 2022). Together, these and our study highlight the relevance of context and urge future clinical trials to further elucidate whether clinical efficacy can be augmented when OT administration is paired with targeted behavioral interventions that support similar states and (social) behaviors.

Another notable observation was the identification that the **parent’s belief about allocated treatment** constituted a potentially important moderator of treatment response, indicating that – within the OT group - parents who believed their child had been assigned the active treatment reported greater benefit than those who believed their child received placebo. Notably, the modulation was only significant in the group receiving the actual OT treatment, not in the placebo group, and only for the immediate post-treatment outcome assessment, not for the four-week follow-up assessment. These results therefore only partly concur with a prior negative OT trial in which moderator effects by parent belief were evident, both in the actual OT group, as well as in the placebo group (Guastella et al., 2015). One the one hand, the modulation by parents’ belief may reflect an expectancy bias, as noted in many prior studies, especially in pediatric trials (King et al., 2009). Particularly in relation to OT intervention research, increased biases can be expected, considering the large media coverage and hype about purported prosocial benefits of the OT “love hormone” that may eventually impact parents’ expectancies (Guastella et al., 2015). However, a sole effect of expectancy bias may be unlikely, since in that case, one would expect response modulations to be present both in the OT and placebo groups. Since the modulating effect was specific to the OT group, the possibility cannot be ruled out that parents may have actually correctly identified real treatment responders, yielding maximal treatment responses in a particular subgroup of children that displayed actual beneficial effects. Further, in line with the notion that context may constitute an important moderator of treatment, one could also envisage that parents who believed their child to receive the active treatment, may have provided their children with more active socio-interactive family contexts during the four-week treatment period, i.e., prompting them to increasingly engage in social experiences and learning, thereby effectively boosting treatment responses.

Another important result relates to the observation that children who **crossed over** from placebo (in phase I) to the actual OT treatment (in phase II), showed a significant further improvement in social responsiveness over and above the substantial placebo-induced improvement noted in phase I. Trial designs in which a phase of blinded placebo intervention is administered before actual treatment allocation have been put forward before as an efficient method to control for placebo effects and to improve detection of ‘real’ therapeutic responses (Yatawara et al., 2016). The current observation of a significant further improvement from a blinded placebo phase (double-blinded) to the active treatment (single-blinded) provides support to this notion. While not explicitly addressed in the current study, it is also noteworthy to stipulate that the oxytocinergic system itself has been suggested to form a key mediator for facilitating placebo-induced improvements. Indeed, as postulated in the recent oxytocin-placebo account of Itskovich et al. (2022); OT is suggested to mediate social facilitation of placebo effects, an effect that is thought to be facilitated by increased social connectedness between patient and clinician during trial participation. In line with this notion, a prior OT administration trial showed that placebo-induced improvements in social functioning coincided with endogenous increases in oxytocin secretion (Parker et al., 2017)

Further, in our trial, children who received the actual OT treatment in the first phase and crossed over to a second phase of active treatment, showed only non-significant within-subject improvements from phase I to phase II, particularly at the four-week follow-up session of phase II - supporting prior observations of a retention of OT’s beneficial effects, also after cessation of the daily nasal spray administrations (Alaerts et al., 2020; Bernaerts et al., 2020). It is noted indeed, that at the last follow-up session of the trial, the majority of children of both the OT-first group and the placebo-first group displayed (reliable) beneficial effects in social responsiveness, indicating that both an eight-week (with a four-week break in the middle) or a continual four-week OT treatment were similarly able to induce a significant beneficial outcome on a core ASD symptom domain. This observation adds to the field’s uncertainty regarding to-be-administered dosing schemas and durations. In multiple-dose OT trials with individuals with ASD, daily dosing ranged from 8-80 IU and durations from 4 continual days to 24 weeks, but strong empirical support for favoring one dosing scheme over another is currently lacking. Some earlier single-dose trials suggested dose-response curves to exhibit U-shaped forms (Lieberz et al., 2020; Spengler et al., 2017), a notion that is supported by a recent chronic four-week OT administration trial in ASD, identifying a daily total dose of 6 IU of TTA-121 (a new formulation of intranasal OT spray with an enhanced bioavailability) to be the most efficacious one, compared to a lower (3 IU) or higher (10 IU) daily dose (Yamasue et al., 2022). Furthermore, in terms of dosing scheme, recent work showed that intermittent (every other day) administration may be therapeutically more efficient than continual administration to obtain anxiolytic effects and reduce amygdala reactivity (Kou et al., 2020). These observations were attributed to reflect a desensitization of the endogenous oxytocinergic system upon too high concentrations and/or too high frequencies of exogenous OT administration. The current observation that a single four-week course can yield the same beneficial effects as a twice four-week course therefore reinforces the notion that longer treatment durations do not necessarily facilitate higher treatment responses. Similarly, in a recent large-scale trial administering OT over a 24-week period, it was noted that the long duration might have attenuated initial early responses to OT (Sikich et al., 2021). In light of these observations, future trials should be directed at identifying the optimal dosing, administration length, and intervals of intranasal OT administration.

To conclude, while the current study showed no overall treatment-specific improvements, important moderator effects were identified, providing initial indications that clinical efficacy can be augmented when OT administration is paired with targeted behavioral interventions that support similar states and (social) behaviors. Future trials are urged to further elucidate the potential of embedding OT treatment within a (socially) stimulating context. Also, the role of parental belief in modulating OT treatment responses needs further attention in subsequent clinical trials. Finally, the observation that (reliable) improvements were established in a large subset of children at the last session of the trial, either after a single four-week course or after two four-week courses, should stimulate future trials to further identify optimal dosing schemas, not only in terms of (daily) concentration, but also in terms of administration length and the possibility of intermittent dosing.

## Data Availability

The data that support the findings of this study are available on reasonable request from the corresponding author, KA. The data are not publicly available due to privacy restrictions.

## Acknowledgements

We would like to thank all the participants of the study and our colleagues of the Leuven Autism Research Consortium (LAuRes).

