## Supplementary Table 1 for "Effects of multiple-dose intranasal oxytocin treatment on social responsiveness in children with autism: A randomized, placebo-controlled trial"

**Full list of inclusion and exclusion criteria.**

| Inclusion criteria | Exclusion criteria |
|---|---|
| Diagnosed with ASD by a multidisciplinary team of experienced clinicians as defined by the DSM-IV-TR or DSM-IV-TR criteria (Diagnostic and Statistical Manual of Mental Disorders). | History of any neurological disorder (stroke, concussion, epilepsy etc). |
| Age-range of 8 to 12 years old. | Significant hearing or vision impairments. |
| Premenstrual girls (girls with onset of menstruation during the course of the trial are allowed to continue the treatment). | Active medical problems: unstable seizures, significant physical illness (e.g., serious liver, renal, or cardiac pathology). |
| Intelligence Quotient above 70 (either full-scaled IQ, verbal IQ or performance IQ). | Regular nasal obstruction or nosebleeds. |
| Dutch native speaker. | Subjects who have had previous chronic treatment with oxytocin. |
| Stable background treatment during four weeks prior to screening. | Participation in another Clinical Trial. |
| No planned changes in psychosocial interventions during the trial. | Known hypersensitivity to active substance or excipients in nasal sprays. |
| | (Significant) change in background treatments. |
| | For MR assessment: any contraindication to MRI research (pacemaker, implanted defibrillator, ear implant / a cochlear implant, insulin or implanted pump, a neurostimulator or VP shunt, any metallic object in the eyes (metallic fragments)*. |

*As indicated in the registration at the European Clinical Trial Registry (Eudract 2018-000769-35), behavioral data collections were part of a larger project assessing (MRI) neurophysiology and biological outcomes.
