## Supplementary Table 2 for "Effects of multiple-dose intranasal oxytocin treatment on social responsiveness in children with autism: A randomized, placebo-controlled trial"

**Detailed information on medication use and comorbidities for participants of the oxytocin and placebo treatment groups.**

Current psychoactive medication use was defined as use within four weeks before study enrollment. Comorbidities were screened through parent-report (with the explicit mentioning of examples in the screening interview including e.g., ADHD, depression, dyscalculia, dyslexia).

All participants had a stable background treatment for at least four weeks prior to the treatment allocation and changes in medication regime were screened and logged.

| | Oxytocin (n = 38) | | Placebo (n = 39) | | Pearson Chi square | *p*-value |
|---|---|---|---|---|---|---|
| | no. | (%) | no. | (%) | | |
| **Psychoactive Medication** | | | | | | |
| Antianginal agents | 1 | (2.5%) | 0 | (0%) | 1.04 | 0.31 |
| Anticholinergic agent | 3 | (7.5%) | 0 | (0%) | 3.20 | 0.07 |
| Anti-depressants | 3 | (7.5%) | 1 | (2.5%) | 1.11 | 0.29 |
| Antipsychotics | 6 | (15%) | 7 | (17.5%) | 0.06 | 0.80 |
| Sleep Aids | 6 | (15%) | 12 | (30%) | 2.41 | 0.12 |
| Stimulants | 11 | (27.5%) | 9 | (22.5%) | 0.34 | 0.56 |
| **Other Medication** | | | | | | |
| Allergy and asthma medications | 3 | (7.5%) | 1 | (2.5%) | 1.11 | 0.29 |
| Gastrointestinal medications | 0 | (0%) | 2 | (5%) | 2.00 | 0.16 |
| Nutritional Supplements | 2 | (5%) | 4 | (10%) | 0.67 | 0.41 |
| Statins | 1 | (2.5%) | 0 | (0%) | 1.04 | 0.31 |
| **Comorbidity** | | | | | | |
| ADHD | 11 | (28.9%) | 10 | (25.6%) | 0.11 | 0.74 |
| DCD | 3 | (7.5%) | 1 | (2.5%) | 1.11 | 0.29 |
| Dyslexia | 2 | (5%) | 5 | (12.5%) | 1.33 | 0.25 |
| Dysorthography | 0 | (0%) | 1 | (2.5%) | 0.99 | 0.32 |
| OCD | 1 | (2.5%) | 0 | (0%) | 1.04 | 0.31 |

ADHD, Attention Deficit Hyperactivity Disorder; DCD, Developmental Coordination Disorder; OCD, Obsessive-Compulsive Disorder.
