## Supplementary Table 3 for "Effects of multiple-dose intranasal oxytocin treatment on social responsiveness in children with autism: A randomized, placebo-controlled trial"

### Side effect screening.

Participants (with the help of their parents) were asked to administer the nasal spray (oxytocin or placebo) daily for four consecutive weeks in phase I (double-blind phase) and another four weeks in phase II (single-blind extension phase). At the end of each week, parents were asked to report whether their child presented any of the listed (or other) side effects and to indicate the severity of the side effect (mild, moderate, or severe). Safety analyses included all participants that received the allocated intervention.

**Panel A** lists the proportion of oxytocin or placebo participants (%) that reported any mild, moderate or severe side effects (averaged across effects). *P*-values correspond to hypothesis tests for difference in proportions between the two treatment groups. Data printed in bold show *p*-values less than 0.05.

**Panel B** lists, separately for each side effect, the proportion of oxytocin (OT) or placebo (PL) participants that reported the side effect, averaged across severity level (mild, moderate, severe).

**Panel C** lists any other incidental adverse event, spontaneously reported by the parents.

While on average, no group differences were evident in the total proportion of reported side effects, a group differences was observed in the proportion of participants who reported moderate side effects in the last week of administration, indicating that the participants of the oxytocin group reported a slightly higher number of moderate side effects during that week.

When examined separately for each side effect, a higher proportion of participants of the oxytocin group reported to experience a ‘headache’ or ‘abdominal/stomach pain’ during the third week of the phase I treatment. Also a higher proportion of participants of the placebo group were noted to experience a ‘sore throat’ and to feel ‘more confident’, compared to the oxytocin group, but only during the first week of the phase I treatment.

In the single-blind phase (phase II), no significant group differences were revealed, either in the total proportion of reported side effects (**Panel A**) or separately for each side effect (**Panel B**).

### Panel A

**Phase I (double-blind)**

| | Oxytocin (%) | | | Placebo (%) | | | Group difference (*p*-value) | | |
|---|---|---|---|---|---|---|---|---|---|
| | Mild | Moderate | Severe | Mild | Moderate | Severe | Mild | Moderate | Severe |
| **Week 1** | 30.00 | 27.50 | 2.50 | 32.50 | 15.00 | 0.00 | 0.81 | 0.17 | 0.31 |
| **Week 2** | 37.50 | 25.00 | 2.50 | 22.50 | 17.50 | 0.00 | 0.14 | 0.41 | 0.31 |
| **Week 3** | 22.50 | 20.00 | 5.00 | 22.50 | 10.00 | 5.00 | 1.00 | 0.21 | 1.00 |
| **Week 4** | 35.00 | 20.00 | 2.50 | 30.00 | 5.00 | 2.50 | 0.63 | **0.04** | 1.00 |
| **Across weeks** | 31.25 | 23.13 | 3.13 | 26.88 | 11.88 | 1.88 | 0.67 | 0.19 | 0.72 |

**Phase II (single-blind)**

| | $\text{Oxytocin}_{\text{first}}$ (%) | | | $\text{Placebo}_{\text{first}}$ (%) | | | Group difference (*p*-value) | | |
|---|---|---|---|---|---|---|---|---|---|
| | Mild | Moderate | Severe | Mild | Moderate | Severe | Mild | Moderate | Severe |
| **Week 1** | 34.21 | 21.05 | 7.89 | 23.08 | 23.08 | 10.26 | 0.28 | 0.83 | 0.72 |
| **Week 2** | 28.95 | 23.68 | 13.16 | 17.95 | 17.95 | 2.56 | 0.25 | 0.54 | 0.08 |
| **Week 3** | 23.68 | 13.16 | 5.26 | 17.95 | 7.69 | 2.56 | 0.54 | 0.43 | 0.54 |
| **Week 4** | 34.21 | 26.32 | 2.63 | 35.90 | 28.21 | 0.00 | 0.88 | 0.85 | 0.31 |
| **Across weeks** | 30.26 | 21.05 | 7.24 | 23.72 | 19.23 | 3.85 | 0.52 | 0.84 | 0.51 |

### Panel B

**Phase I (double-blind)**
**(OT *n* = 40; PL *n* = 40)**

| | Week 1 | | | Week 2 | | | Week 3 | | | Week 4 | | |
|---|---|---|---|---|---|---|---|---|---|---|---|---|
| | OT (%) | PL (%) | *p* | OT (%) | PL (%) | *p* | OT (%) | PL (%) | *p* | OT (%) | PL (%) | *p* |
| Headache | 10.00 | 12.50 | 0.72 | 7.50 | 7.50 | 1.00 | 12.50 | 0.00 | **0.02** | 5.00 | 2.50 | 0.56 |
| Drowsiness | 2.50 | 2.50 | 1.00 | 7.50 | 5.00 | 0.64 | 7.50 | 0.00 | 0.08 | 2.50 | 0.00 | 0.31 |
| Dizziness | 7.50 | 2.50 | 0.30 | 2.50 | 2.50 | 1.00 | 7.50 | 0.00 | 0.08 | 2.50 | 0.00 | 0.31 |
| Fainting | 0.00 | 0.00 | 1.00 | 0.00 | 0.00 | 1.00 | 2.50 | 0.00 | 0.31 | 0.00 | 0.00 | 1.00 |
| Changes in heart rate or palpitations | 0.00 | 0.00 | 1.00 | 2.50 | 0.00 | 0.31 | 2.50 | 0.00 | 0.31 | 0.00 | 0.00 | 1.00 |
| Shortness of breath | 0.00 | 0.00 | 1.00 | 0.00 | 0.00 | 1.00 | 0.00 | 2.50 | 0.31 | 0.00 | 0.00 | 1.00 |
| Fever | 0.00 | 2.50 | 0.31 | 0.00 | 0.00 | 1.00 | 0.00 | 0.00 | 1.00 | 2.50 | 0.00 | 0.31 |
| Sore throat | 0.00 | 12.50 | **0.02** | 5.00 | 7.50 | 0.64 | 5.00 | 0.00 | 0.15 | 0.00 | 0.00 | 1.00 |
| Dry throat/dry mouth | 0.00 | 5.00 | 0.15 | 0.00 | 0.00 | 1.00 | 2.50 | 0.00 | 0.31 | 5.00 | 0.00 | 0.15 |
| Hoarseness | 0.00 | 5.00 | 0.15 | 0.00 | 2.50 | 0.31 | 0.00 | 0.00 | 1.00 | 2.50 | 0.00 | 0.31 |
| Coughing | 5.00 | 5.00 | 1.00 | 2.50 | 2.50 | 1.00 | 2.50 | 0.00 | 0.31 | 5.00 | 0.00 | 0.15 |
| Coughing up mucus | 0.00 | 0.00 | 1.00 | 0.00 | 0.00 | 1.00 | 0.00 | 0.00 | 1.00 | 0.00 | 0.00 | 1.00 |
| Congested nose | 10.00 | 7.50 | 0.69 | 5.00 | 10.00 | 0.40 | 10.00 | 5.00 | 0.40 | 5.00 | 5.00 | 1.00 |
| Sneezing | 2.50 | 2.50 | 1.00 | 0.00 | 2.50 | 0.31 | 7.50 | 2.50 | 0.30 | 7.50 | 2.50 | 0.30 |
| Nasal irritation | 5.00 | 5.00 | 1.00 | 5.00 | 0.00 | 0.15 | 7.50 | 2.50 | 0.30 | 2.50 | 0.00 | 0.31 |
| Runny nose | 2.50 | 2.50 | 1.00 | 2.50 | 2.50 | 1.00 | 2.50 | 0.00 | 0.31 | 7.50 | 0.00 | 0.08 |
| Burning sensation in nose and/or ears | 5.00 | 0.00 | 0.15 | 5.00 | 0.00 | 0.15 | 5.00 | 0.00 | 0.15 | 2.50 | 0.00 | 0.31 |
| Sensitive to fragrances | 0.00 | 0.00 | 1.00 | 0.00 | 0.00 | 1.00 | 0.00 | 0.00 | 1.00 | 2.50 | 0.00 | 0.31 |
| Watery eyes | 0.00 | 0.00 | 1.00 | 2.50 | 0.00 | 0.31 | 0.00 | 0.00 | 1.00 | 2.50 | 0.00 | 0.31 |

| | | | | | | | | | | | | |
|---|---|---|---|---|---|---|---|---|---|---|---|---|
| Nausea and/or vomiting | 2.50 | 7.50 | 0.30 | 5.00 | 2.50 | 0.56 | 2.50 | 0.00 | 0.31 | 0.00 | 0.00 | 1.00 |
| Abdominal or stomach pain | 12.50 | 10.00 | 0.72 | 15.00 | 7.50 | 0.29 | 12.50 | 0.00 | **0.02** | 7.50 | 7.50 | 1.00 |
| Decreased appetite | 2.50 | 5.00 | 0.56 | 2.50 | 2.50 | 1.00 | 5.00 | 0.00 | 0.15 | 2.50 | 0.00 | 0.31 |
| Hungry or increased appetite | 0.00 | 2.50 | 0.31 | 0.00 | 2.50 | 0.31 | 2.50 | 0.00 | 0.31 | 0.00 | 0.00 | 1.00 |
| Constipation | 7.50 | 2.50 | 0.30 | 5.00 | 0.00 | 0.15 | 7.50 | 0.00 | 0.08 | 7.50 | 2.50 | 0.30 |
| Diarrhea | 0.00 | 2.50 | 0.31 | 2.50 | 2.50 | 1.00 | 2.50 | 0.00 | 0.31 | 0.00 | 0.00 | 1.00 |
| Muscle pain/cramps | 5.00 | 0.00 | 0.15 | 5.00 | 0.00 | 0.15 | 5.00 | 0.00 | 0.15 | 5.00 | 2.50 | 0.56 |
| Skin rash | 2.50 | 0.00 | 0.31 | 2.50 | 0.00 | 0.31 | 0.00 | 0.00 | 1.00 | 5.00 | 0.00 | 0.15 |
| Increased fluid intake | 0.00 | 0.00 | 1.00 | 0.00 | 0.00 | 1.00 | 2.50 | 0.00 | 0.31 | 0.00 | 0.00 | 1.00 |
| Water retention/bloating | 0.00 | 0.00 | 1.00 | 2.50 | 0.00 | 0.31 | 0.00 | 0.00 | 1.00 | 2.50 | 0.00 | 0.31 |
| Insomnia/sleep difficulties | 5.00 | 0.00 | 0.15 | 5.00 | 5.00 | 1.00 | 5.00 | 0.00 | 0.15 | 0.00 | 5.00 | 0.15 |
| Nightmares | 2.50 | 0.00 | 0.31 | 0.00 | 0.00 | 1.00 | 0.00 | 0.00 | 1.00 | 0.00 | 0.00 | 1.00 |
| Staring/daydreams | 0.00 | 0.00 | 1.00 | 2.50 | 0.00 | 0.31 | 0.00 | 0.00 | 1.00 | 2.50 | 0.00 | 0.31 |
| Anaphylaxis | 0.00 | 0.00 | 1.00 | 0.00 | 0.00 | 1.00 | 0.00 | 0.00 | 1.00 | 0.00 | 0.00 | 1.00 |
| Changes in perception of the tongue | 0.00 | 0.00 | 1.00 | 0.00 | 0.00 | 1.00 | 0.00 | 0.00 | 1.00 | 0.00 | 0.00 | 1.00 |
| Back pain | 0.00 | 0.00 | 1.00 | 0.00 | 0.00 | 1.00 | 0.00 | 0.00 | 1.00 | 0.00 | 0.00 | 1.00 |
| Bed wetting | 0.00 | 2.50 | 0.31 | 0.00 | 5.00 | 0.15 | 0.00 | 0.00 | 1.00 | 0.00 | 0.00 | 1.00 |
| Weight gain | 0.00 | 0.00 | 1.00 | 0.00 | 0.00 | 1.00 | 0.00 | 0.00 | 1.00 | 0.00 | 0.00 | 1.00 |
| Sweating | 0.00 | 0.00 | 1.00 | 2.50 | 0.00 | 0.31 | 0.00 | 0.00 | 1.00 | 0.00 | 0.00 | 1.00 |
| Blurred vision | 0.00 | 0.00 | 1.00 | 0.00 | 0.00 | 1.00 | 0.00 | 0.00 | 1.00 | 0.00 | 0.00 | 1.00 |
| Less talk to others | 0.00 | 0.00 | 1.00 | 0.00 | 2.50 | 0.31 | 2.50 | 0.00 | 0.31 | 2.50 | 2.50 | 1.00 |
| Uninterested in others | 0.00 | 0.00 | 1.00 | 0.00 | 2.50 | 0.31 | 0.00 | 0.00 | 1.00 | 2.50 | 0.00 | 0.31 |
| Persistent thoughts and/or feelings | 2.50 | 7.50 | 0.30 | 0.00 | 5.00 | 0.15 | 2.50 | 2.50 | 1.00 | 0.00 | 2.50 | 0.31 |
| Development of repetitive behavior | 0.00 | 0.00 | 1.00 | 2.50 | 0.00 | 0.31 | 0.00 | 2.50 | 0.31 | 0.00 | 0.00 | 1.00 |
| Increase in repetitive behavior | 2.50 | 0.00 | 0.31 | 2.50 | 0.00 | 0.31 | 0.00 | 2.50 | 0.31 | 0.00 | 0.00 | 1.00 |
| Nail biting | 0.00 | 0.00 | 1.00 | 0.00 | 0.00 | 1.00 | 0.00 | 0.00 | 1.00 | 2.50 | 0.00 | 0.31 |
| Irritability or Anger | 0.00 | 7.50 | 0.08 | 2.50 | 7.50 | 0.30 | 7.50 | 5.00 | 0.64 | 10.00 | 7.50 | 0.69 |
| Sad | 2.50 | 0.00 | 0.31 | 0.00 | 7.50 | 0.08 | 2.50 | 5.00 | 0.56 | 5.00 | 5.00 | 1.00 |
| Prone to crying or more emotional | 2.50 | 5.00 | 0.56 | 2.50 | 12.50 | 0.09 | 7.50 | 12.50 | 0.46 | 5.00 | 7.50 | 0.64 |
| Anxious, worried or discomfort | 0.00 | 0.00 | 1.00 | 5.00 | 0.00 | 0.15 | 5.00 | 2.50 | 0.56 | 2.50 | 2.50 | 1.00 |
| Happy or satisfied | 10.00 | 25.00 | 0.08 | 7.50 | 15.00 | 0.29 | 17.50 | 15.00 | 0.76 | 7.50 | 12.50 | 0.46 |
| Euphoric or unusually happy | 5.00 | 10.00 | 0.40 | 7.50 | 7.50 | 1.00 | 7.50 | 5.00 | 0.64 | 10.00 | 2.50 | 0.17 |
| Calm, relaxed or comfortable | 10.00 | 25.00 | 0.08 | 15.00 | 17.50 | 0.76 | 12.50 | 22.50 | 0.24 | 7.50 | 10.00 | 0.69 |
| More focused | 0.00 | 2.50 | 0.31 | 0.00 | 5.00 | 0.15 | 2.50 | 2.50 | 1.00 | 0.00 | 2.50 | 0.31 |
| More confidence | 0.00 | 12.50 | **0.02** | 7.50 | 7.50 | 1.00 | 7.50 | 5.00 | 0.64 | 5.00 | 7.50 | 0.64 |

| | **Phase II (single-blind) (OT$_{first}$ *n* = 38; PL$_{first}$ *n* = 39)** | | | | | | | | | | | |
|---|---|---|---|---|---|---|---|---|---|---|---|---|
| | **Week 1** | | | **Week 2** | | | **Week 3** | | | **Week 4** | | |
| | **OT$_{first}$ (%)** | **PL$_{first}$ (%)** | ***p*** | **OT$_{first}$ (%)** | **PL$_{first}$ (%)** | ***p*** | **OT$_{first}$ (%)** | **PL$_{first}$ (%)** | ***p*** | **OT$_{first}$ (%)** | **PL$_{first}$ (%)** | ***P*** |
| Headache | 5.26 | 10.26 | 0.41 | 2.63 | 7.69 | 0.32 | 10.53 | 5.13 | 0.38 | 5.26 | 2.56 | 0.54 |
| Drowsiness | 2.63 | 0.00 | 0.31 | 2.63 | 2.56 | 0.99 | 0.00 | 0.00 | 1.00 | 0.00 | 0.00 | 1.00 |
| Dizziness | 0.00 | 2.56 | 0.32 | 0.00 | 2.56 | 0.32 | 0.00 | 0.00 | 1.00 | 0.00 | 0.00 | 1.00 |
| Fainting | 0.00 | 0.00 | 1.00 | 0.00 | 0.00 | 1.00 | 0.00 | 0.00 | 1.00 | 0.00 | 0.00 | 1.00 |
| Changes in heart rate or palpitations | 0.00 | 0.00 | 1.00 | 0.00 | 0.00 | 1.00 | 0.00 | 0.00 | 1.00 | 5.26 | 0.00 | 0.15 |
| Shortness of breath | 0.00 | 0.00 | 1.00 | 0.00 | 0.00 | 1.00 | 0.00 | 2.56 | 0.32 | 0.00 | 0.00 | 1.00 |

| | | | | | | | | | | | | |
|---|---|---|---|---|---|---|---|---|---|---|---|---|
| Fever | 0.00 | 2.56 | 0.32 | 0.00 | 2.56 | 0.32 | 0.00 | 0.00 | 1.00 | 0.00 | 2.56 | 0.32 |
| Sore throat | 2.63 | 5.13 | 0.57 | 2.63 | 7.69 | 0.32 | 5.26 | 5.13 | 0.98 | 5.26 | 0.00 | 0.15 |
| Dry throat/dry mouth | 0.00 | 2.56 | 0.32 | 7.89 | 2.56 | 0.29 | 5.26 | 2.56 | 0.54 | 5.26 | 2.56 | 0.54 |
| Hoarseness | 0.00 | 0.00 | 1.00 | 2.63 | 0.00 | 0.31 | 2.63 | 2.56 | 0.99 | 2.63 | 0.00 | 0.31 |
| Coughing | 2.63 | 2.56 | 0.99 | 2.63 | 0.00 | 0.31 | 2.63 | 2.56 | 0.99 | 5.26 | 0.00 | 0.15 |
| Coughing up mucus | 0.00 | 0.00 | 1.00 | 2.63 | 0.00 | 0.31 | 2.63 | 0.00 | 0.31 | 0.00 | 0.00 | 1.00 |
| Congested nose | 2.63 | 5.13 | 0.57 | 5.26 | 5.13 | 0.98 | 5.26 | 7.69 | 0.67 | 7.89 | 0.00 | 0.07 |
| Sneezing | 0.00 | 0.00 | 1.00 | 0.00 | 2.56 | 0.32 | 0.00 | 2.56 | 0.32 | 2.63 | 0.00 | 0.31 |
| Nasal irritation | 2.63 | 5.13 | 0.57 | 0.00 | 0.00 | 1.00 | 5.26 | 2.56 | 0.54 | 7.89 | 0.00 | 0.07 |
| Runny nose | 2.63 | 0.00 | 0.31 | 0.00 | 2.56 | 0.32 | 2.63 | 5.13 | 0.57 | 0.00 | 5.13 | 0.16 |
| Burning sensation in nose and/or ears | 0.00 | 2.56 | 0.32 | 0.00 | 0.00 | 1.00 | 2.63 | 0.00 | 0.31 | 0.00 | 0.00 | 1.00 |
| Sensitive to fragrances | 0.00 | 0.00 | 1.00 | 2.63 | 2.56 | 0.99 | 2.63 | 0.00 | 0.31 | 2.63 | 0.00 | 0.31 |
| Watery eyes | 0.00 | 0.00 | 1.00 | 0.00 | 0.00 | 1.00 | 2.63 | 0.00 | 0.31 | 2.63 | 0.00 | 0.31 |
| Nausea and/or vomiting | 2.63 | 0.00 | 0.31 | 5.26 | 0.00 | 0.15 | 2.63 | 2.56 | 0.99 | 2.63 | 0.00 | 0.31 |
| Abdominal or stomach pain | 5.26 | 2.56 | 0.54 | 13.16 | 7.69 | 0.43 | 5.26 | 2.56 | 0.54 | 2.63 | 2.56 | 0.99 |
| Decreased appetite | 2.63 | 0.00 | 0.31 | 7.89 | 7.69 | 0.97 | 2.63 | 5.13 | 0.57 | 2.63 | 2.56 | 0.99 |
| Hungry or increased appetite | 0.00 | 0.00 | 1.00 | 2.63 | 2.56 | 0.99 | 0.00 | 0.00 | 1.00 | 0.00 | 0.00 | 1.00 |
| Constipation | 0.00 | 0.00 | 1.00 | 2.63 | 0.00 | 0.31 | 0.00 | 0.00 | 1.00 | 0.00 | 0.00 | 1.00 |
| Diarrhea | 5.26 | 0.00 | 0.15 | 5.26 | 0.00 | 0.15 | 0.00 | 0.00 | 1.00 | 0.00 | 0.00 | 1.00 |
| Muscle pain/cramps | 7.89 | 0.00 | 0.07 | 2.63 | 5.13 | 0.57 | 2.63 | 0.00 | 0.31 | 5.26 | 0.00 | 0.15 |
| Skin rash | 2.63 | 0.00 | 0.31 | 5.26 | 0.00 | 0.15 | 2.63 | 0.00 | 0.31 | 0.00 | 2.56 | 0.32 |
| Increased fluid intake | 0.00 | 2.56 | 0.32 | 2.63 | 0.00 | 0.31 | 5.26 | 2.56 | 0.54 | 2.63 | 0.00 | 0.31 |
| Water retention/bloating | 0.00 | 0.00 | 1.00 | 0.00 | 0.00 | 1.00 | 0.00 | 0.00 | 1.00 | 0.00 | 0.00 | 1.00 |
| Insomnia/sleep difficulties | 0.00 | 0.00 | 1.00 | 0.00 | 2.56 | 0.32 | 2.63 | 2.56 | 0.99 | 5.26 | 2.56 | 0.54 |
| Nightmares | 2.63 | 0.00 | 0.31 | 2.63 | 0.00 | 0.31 | 0.00 | 0.00 | 1.00 | 0.00 | 0.00 | 1.00 |
| Staring/daydreams | 2.63 | 0.00 | 0.31 | 2.63 | 2.56 | 0.99 | 0.00 | 0.00 | 1.00 | 0.00 | 0.00 | 1.00 |
| Anaphylaxis | 0.00 | 0.00 | 1.00 | 0.00 | 0.00 | 1.00 | 0.00 | 0.00 | 1.00 | 0.00 | 0.00 | 1.00 |
| Changes in perception of the tongue | 0.00 | 0.00 | 1.00 | 0.00 | 0.00 | 1.00 | 0.00 | 0.00 | 1.00 | 0.00 | 0.00 | 1.00 |
| Back pain | 0.00 | 0.00 | 1.00 | 0.00 | 0.00 | 1.00 | 0.00 | 0.00 | 1.00 | 0.00 | 0.00 | 1.00 |
| Bed wetting | 0.00 | 0.00 | 1.00 | 2.63 | 0.00 | 0.31 | 0.00 | 0.00 | 1.00 | 0.00 | 0.00 | 1.00 |
| Weight gain | 0.00 | 0.00 | 1.00 | 0.00 | 2.56 | 0.32 | 0.00 | 0.00 | 1.00 | 0.00 | 0.00 | 1.00 |
| Sweating | 0.00 | 0.00 | 1.00 | 2.63 | 0.00 | 0.31 | 2.63 | 2.56 | 0.99 | 2.63 | 2.56 | 0.99 |
| Blurred vision | 0.00 | 0.00 | 1.00 | 0.00 | 0.00 | 1.00 | 0.00 | 0.00 | 1.00 | 0.00 | 0.00 | 1.00 |
| Less talk to others | 0.00 | 0.00 | 1.00 | 0.00 | 7.69 | 0.08 | 0.00 | 2.56 | 0.32 | 2.63 | 2.56 | 0.99 |
| Uninterested in others | 0.00 | 0.00 | 1.00 | 0.00 | 2.56 | 0.32 | 2.63 | 0.00 | 0.31 | 2.63 | 0.00 | 0.31 |
| Persistent thoughts and/or feelings | 0.00 | 2.56 | 0.32 | 2.63 | 5.13 | 0.57 | 2.63 | 0.00 | 0.31 | 5.26 | 0.00 | 0.15 |
| Development of repetitive behavior | 5.26 | 0.00 | 0.15 | 0.00 | 0.00 | 1.00 | 0.00 | 2.56 | 0.32 | 0.00 | 0.00 | 1.00 |
| Increase in repetitive behavior | 0.00 | 0.00 | 1.00 | 2.63 | 2.56 | 0.99 | 0.00 | 0.00 | 1.00 | 0.00 | 0.00 | 1.00 |
| Nail biting | 2.63 | 0.00 | 0.31 | 2.63 | 0.00 | 0.31 | 0.00 | 0.00 | 1.00 | 0.00 | 2.56 | 0.32 |
| Irritability or Anger | 2.63 | 2.56 | 0.99 | 7.89 | 10.26 | 0.72 | 5.26 | 5.13 | 0.98 | 5.26 | 0.00 | 0.15 |
| Sad | 2.63 | 0.00 | 0.31 | 7.89 | 5.13 | 0.62 | 2.63 | 0.00 | 0.31 | 0.00 | 0.00 | 1.00 |
| Prone to crying or more emotional | 2.63 | 2.56 | 0.99 | 13.16 | 2.56 | 0.08 | 7.89 | 0.00 | 0.07 | 2.63 | 5.13 | 0.57 |
| Anxious, worried or discomfort | 2.63 | 0.00 | 0.31 | 5.26 | 2.56 | 0.54 | 5.26 | 0.00 | 0.15 | 2.63 | 0.00 | 0.31 |
| Happy or satisfied | 21.05 | 12.82 | 0.33 | 15.79 | 12.82 | 0.71 | 7.89 | 7.69 | 0.97 | 5.26 | 7.69 | 0.67 |
| Euphoric or unusually happy | 5.26 | 2.56 | 0.54 | 5.26 | 0.00 | 0.15 | 2.63 | 2.56 | 0.99 | 2.63 | 0.00 | 0.31 |
| Calm, relaxed or | 18.42 | 15.38 | 0.72 | 13.16 | 12.82 | 0.96 | 10.53 | 10.26 | 0.97 | 7.89 | 12.82 | 0.48 |

| | | | | | | | | | | | | |
|---|---|---|---|---|---|---|---|---|---|---|---|---|
| comfortable | | | | | | | | | | | | |
| More focused | 5.26 | 0.00 | 0.15 | 5.26 | 5.13 | 0.98 | 0.00 | 5.13 | 0.16 | 0.00 | 0.00 | 1.00 |
| More confidence | 7.89 | 5.13 | 0.62 | 7.89 | 5.13 | 0.62 | 0.00 | 7.69 | 0.08 | 0.00 | 7.69 | 0.08 |

| **Panel C** | **Phase I (double-blind)** | | | | | | | | **Phase II (single-blind)** | | | | | | | |
|---|---|---|---|---|---|---|---|---|---|---|---|---|---|---|---|---|
| **Number of other incidental events in spontaneous reports** | **Week 1** | | **Week 2** | | **Week 3** | | **Week 4** | | **Week 1** | | **Week 2** | | **Week 3** | | **Week 4** | |
| | **OT** | **PL** | **OT** | **PL** | **OT** | **PL** | **OT** | **PL** | **$OT_{first}$** | **$PL_{first}$** | **$OT_{first}$** | **$PL_{first}$** | **$OT_{first}$** | **$PL_{first}$** | **$OT_{first}$** | **$PL_{first}$** |
| Nose bleeding | 0 | 0 | 0 | 1 | 0 | 0 | 0 | 1 | 0 | 0 | 0 | 1 | 0 | 0 | 0 | 0 |
| Fracture | 0 | 0 | 0 | 0 | 0 | 0 | 0 | 0 | 0 | 0 | 0 | 0 | 0 | 0 | 0 | 1 |
| Earache | 0 | 0 | 1 | 0 | 0 | 0 | 0 | 0 | 0 | 0 | 0 | 0 | 0 | 0 | 0 | 0 |
| Inappropriate Affect | 0 | 1 | 0 | 0 | 0 | 1 | 0 | 1 | 0 | 1 | 0 | 1 | 1 | 0 | 1 | 1 |
| Mood swings | 0 | 0 | 0 | 0 | 0 | 0 | 1 | 0 | 0 | 0 | 0 | 0 | 0 | 0 | 0 | 0 |
| Less focussed | 1 | 0 | 0 | 0 | 0 | 0 | 1 | 0 | 3 | 0 | 2 | 0 | 0 | 0 | 0 | 0 |
| Fecal incontinence | 0 | 0 | 0 | 0 | 0 | 0 | 1 | 0 | 0 | 0 | 0 | 0 | 0 | 0 | 0 | 0 |
| Feeling cold | 0 | 1 | 0 | 0 | 0 | 0 | 0 | 0 | 0 | 0 | 0 | 0 | 0 | 0 | 0 | 0 |
