## Supplementary Table 4 for "Effects of multiple-dose intranasal oxytocin treatment on social responsiveness in children with autism: A randomized, placebo-controlled trial"

### Daily screenings of changes in affect and arousal.

Both participants and their parents completed a structured daily diary for four consecutive weeks in phase I (double-blind phase) and another four weeks in phase II (single-blind extension phase). Once daily, parents were asked to complete two 9-point Manikin rating scales, rating their child's perceived arousal (1 = calm, 9 = excited) and valence (1 = feeling pleasant/happy, 9 = feeling unpleasant/unhappy; Bradley & Lang; 1994). Children also completed a self-report of the arousal and valence scale, twice-daily, once at noon and once in the evening.

To examine treatment-related differences in the daily screenings, weekly averages were calculated for each rating and subjected to a general linear model with the within-subject factor 'week' (week 1-4) and the between-subject factors 'treatment' (oxytocin, placebo).

None of the scales yielded a significant main effect of 'treatment' **across weeks** (all, $p > .05$), indicating no overall significant group differences in ratings of valence or arousal, either by the parents or by the child, both in phase I and phase II.

Closer analysis of treatment effects **separately for each week** revealed a significant group difference in child-ratings of valence (in the evening), indicating higher feelings of 'unpleasantness' in the oxytocin, compared to the placebo group during the second week of the phase I treatment. Also in the first week of phase II, a similar effect was noted, indicating that children who crossed over from the placebo treatment (in phase I) to the oxytocin treatment (in phase II) (placebo-first group) reported slightly higher feelings of unpleasantness, compared to children who were receiving their second course of oxytocin treatment (oxytocin-first group).

In the table below, weekly averages (and average responses across the four weeks) are reported separately for each treatment group (oxytocin, placebo) and phase (phase I and II). *P*-values correspond to independent-sample *t*-tests (or *F*-tests) assessing between-group differences in ratings of arousal and valence. Data printed in bold show *p*-values less than 0.05.

Notably, for several of the scales, the general linear model also revealed a main effect of week, indicating that across treatment groups, both parents and children reported improvements in reports of valence (more pleasant) and arousal (more calm) from the first to the last week of the trial, particularly in phase II, when all children received the 'actual' oxytocin treatment (phase II, parent-reported valence: $F(3,216) = 4.14$; $p = .007$; child-reported valence (noon & evening): $F(3,213) = 3.09$; $p < .028$; child-reported arousal (noon & evening) $F(3,213) = 6.88$; $p < .001$). Also in phase I, an overall effect of 'week' was evident in terms of child-reported valence (only at noon) ($F(3,195) = 5.62$; $p = .001$).

In addition to the ratings of arousal and valence, parents were also asked to indicate how they experienced the interaction with their child while completing the daily diary together, using a 5-point scale (1 = unpleasant/difficult; 5 = pleasant/easy). Generally, child-parent interactions while completing the daily diaries were experienced to be overall pleasant/easy, with slightly more pleasant experiences of the child-parent interaction in the oxytocin group, compared to the placebo group in phase I of the trial (oxytocin (n = 38): 4.32 ± 0.47; placebo (n = 36): 4.06 ± 0.58; $t(72) = 2.26$, $p = .027$). Differences in the experiences of the child-parent interaction were no longer evident in phase II of the trial, when all children received the actual oxytocin treatment (oxytocin-first (n = 36): 4.14 ± 0.73; placebo-first (n = 37): 3.84 ± 0.84; $t(71) = 1.62$, $p = .11$). Generally, these overall positive ratings provide an indication that the completion of the daily diaries was well-tolerated, both by the children and their parents.

**Reference:** Bradley, M. M., & Lang, P. J. (1994). Measuring emotion: the Self-Assessment Manikin and the Semantic Differential. *Journal of Behavior Therapy and Experimental Psychiatry*, 25(1), 49–59. https://doi.org/10.1016/0005-7916(94)90063-9

| | Phase I (double-blind) | | | | | Phase II (single-blind) | | | | |
|---|---|---|---|---|---|---|---|---|---|---|
| | Oxytocin | | Placebo | | | Oxytocin$_{first}$ | | Placebo$_{first}$ | | |
| | N | Mean ± SD | N | Mean ± SD | *p*-value | N | Mean ± SD | N | Mean ± SD | *p*-value |
| **Week 1** | | | | | | | | | | |
| **Informant report** | | | | | | | | | | |
| Valence | 37 | 3.10 ± 0.92 | 36 | 3.03 ± 1.17 | 0.780 | 36 | 3.74 ± 1.54 | 38 | 3.55 ± 1.23 | 0.565 |
| Arousal | 37 | 3.70 ± 1.13 | 36 | 3.64 ± 1.24 | 0.831 | 36 | 3.05 ± 1.03 | 38 | 3.30 ± 1.36 | 0.366 |
| **Self report** | | | | | | | | | | |
| Valence - noon | 36 | 2.71 ± 1.15 | 35 | 2.68 ± 1.19 | 0.905 | 37 | 2.36 ± 1.11 | 39 | 2.67 ± 1.32 | 0.280 |
| Arousal - noon | 36 | 3.30 ± 1.20 | 35 | 3.20 ± 1.25 | 0.749 | 37 | 3.07 ± 1.51 | 39 | 3.27 ± 1.57 | 0.578 |
| Valence - evening | 36 | 2.69 ± 1.15 | 35 | 2.70 ± 1.13 | 0.957 | 37 | 2.25 ± 1.07 | 39 | 2.92 ± 1.32 | **0.017** |
| Arousal - evening | 36 | 3.47 ± 1.29 | 35 | 3.16 ± 1.07 | 0.279 | 37 | 3.10 ± 1.67 | 39 | 3.49 ± 1.48 | 0.283 |
| **Week 2** | | | | | | | | | | |
| **Informant report** | | | | | | | | | | |
| Valence | 37 | 3.23 ± 1.04 | 36 | 2.96 ± 1.32 | 0.323 | 36 | 3.55 ± 1.55 | 38 | 3.32 ± 1.11 | 0.461 |
| Arousal | 37 | 3.93 ± 1.45 | 36 | 3.55 ± 1.36 | 0.246 | 36 | 2.79 ± 0.96 | 38 | 2.99 ± 1.27 | 0.446 |
| **Self report** | | | | | | | | | | |
| Valence - noon | 36 | 2.78 ± 1.25 | 35 | 2.31 ± 1.20 | 0.109 | 35 | 2.02 ± 0.94 | 38 | 2.41 ± 1.24 | 0.139 |
| Arousal - noon | 36 | 3.58 ± 1.66 | 35 | 3.00 ± 1.53 | 0.135 | 35 | 2.92 ± 1.71 | 38 | 2.89 ± 1.40 | 0.922 |
| Valence - evening | 36 | 3.01 ± 1.28 | 35 | 2.32 ± 1.15 | **0.019** | 35 | 2.32 ± 1.04 | 38 | 2.53 ± 1.21 | 0.441 |
| Arousal - evening | 36 | 3.70 ± 1.49 | 35 | 3.04 ± 1.38 | 0.057 | 35 | 3.20 ± 1.69 | 38 | 2.94 ± 1.37 | 0.477 |
| **Week 3** | | | | | | | | | | |
| **Informant report** | | | | | | | | | | |
| Valence | 37 | 3.19 ± 1.09 | 36 | 2.95 ± 1.08 | 0.345 | 37 | 3.83 ± 1.97 | 39 | 3.23 ± 1.41 | 0.124 |
| Arousal | 37 | 3.86 ± 1.38 | 36 | 3.32 ± 1.31 | 0.092 | 37 | 3.03 ± 1.24 | 39 | 2.80 ± 1.19 | 0.422 |
| **Self report** | | | | | | | | | | |
| Valence - noon | 36 | 2.47 ± 1.15 | 34 | 2.25 ± 1.15 | 0.434 | 36 | 2.03 ± 1.04 | 38 | 2.29 ± 1.08 | 0.286 |
| Arousal - noon | 36 | 3.21 ± 1.47 | 34 | 3.01 ± 1.67 | 0.588 | 36 | 2.80 ± 1.68 | 38 | 2.62 ± 1.37 | 0.618 |
| Valence - evening | 36 | 2.73 ± 1.11 | 34 | 2.39 ± 1.27 | 0.230 | 36 | 2.20 ± 0.99 | 38 | 2.37 ± 1.15 | 0.504 |
| Arousal - evening | 36 | 3.26 ± 1.33 | 34 | 3.17 ± 1.73 | 0.801 | 36 | 2.96 ± 1.77 | 38 | 2.52 ± 1.24 | 0.226 |
| **Week 4** | | | | | | | | | | |
| **Informant report** | | | | | | | | | | |
| Valence | 35 | 3.11 ± 0.81 | 36 | 2.89 ± 1.26 | 0.383 | 37 | 3.63 ± 1.86 | 39 | 3.13 ± 1.33 | 0.179 |
| Arousal | 35 | 3.79 ± 1.35 | 36 | 3.23 ± 1.31 | 0.081 | 37 | 2.89 ± 1.28 | 39 | 2.79 ± 1.15 | 0.724 |
| **Self report** | | | | | | | | | | |
| Valence - noon | 34 | 2.51 ± 1.09 | 34 | 2.23 ± 1.17 | 0.306 | 36 | 2.03 ± 0.98 | 38 | 2.08 ± 0.95 | 0.801 |
| Arousal - noon | 34 | 3.03 ± 1.37 | 34 | 2.91 ± 1.42 | 0.721 | 36 | 2.85 ± 1.74 | 38 | 2.57 ± 1.43 | 0.450 |
| Valence - evening | 34 | 2.72 ± 1.18 | 34 | 2.51 ± 1.25 | 0.467 | 36 | 2.18 ± 1.05 | 38 | 2.34 ± 1.10 | 0.533 |
| Arousal - evening | 34 | 3.35 ± 1.23 | 34 | 3.16 ± 1.65 | 0.607 | 36 | 3.07 ± 1.79 | 38 | 2.79 ± 1.45 | 0.462 |
| **Across weeks** | | | | | | | | | | |
| **Informant report** | | | | | | | | | | |
| Valence | 37 | 3.17 ± 0.70 | 36 | 2.96 ± 1.00 | 0.302 | 36 | 3.69 ± 1.60 | 38 | 3.26 ± 1.00 | 0.163 |
| Arousal | 37 | 3.83 ± 1.15 | 36 | 3.43 ± 1.06 | 0.131 | 36 | 2.94 ± 0.95 | 38 | 2.93 ± 1.05 | 0.969 |
| **Self report** | | | | | | | | | | |
| Valence - noon | 36 | 2.62 ± 0.98 | 35 | 2.36 ± 1.10 | 0.282 | 37 | 2.18 ± 1.02 | 39 | 2.42 ± 1.09 | 0.333 |
| Arousal - noon | 36 | 2.79 ± 1.02 | 35 | 2.46 ± 1.02 | 0.175 | 37 | 2.24 ± 0.86 | 39 | 2.59 ± 1.06 | 0.120 |
| Valence - evening | 36 | 3.29 ± 1.23 | 35 | 3.01 ± 1.29 | 0.355 | 37 | 2.92 ± 1.54 | 39 | 2.88 ± 1.38 | 0.917 |
| Arousal - evening | 36 | 3.45 ± 1.14 | 35 | 3.11 ± 1.28 | 0.247 | 37 | 3.06 ± 1.62 | 39 | 2.99 ± 1.24 | 0.818 |

**Valence scale:** Higher scores denote feeling more unpleasant/unhappy (1 = feeling pleasant/happy, 9 = feeling unpleasant/unhappy). **Arousal scale:** Higher scores denote feeling more excited (1 = calm, 9 = excited).
