## Supplementary Table 5 for "Effects of multiple-dose intranasal oxytocin treatment on social responsiveness in children with autism: A randomized, placebo-controlled trial"

**Detailed description of questionnaires adopted as descriptive, primary or secondary outcomes.**

| Questionnaire | Construct | Type of outcome | Type of report | Number of items | Range of scores | Rating scale (points) | Meaning of higher scores | Reference |
|---|---|---|---|---|---|---|---|---|
| **Autism Diagnostic Observation Schedule (ADOS-2)** | Symptom severity | Descriptive | Observation | Per module | 1-10 | - | More severe symptoms of autism spectrum disorder | Lord et al., 2012 |
| **Wechsler Intelligence Scale for Children (WISC-V-NL)** | Verbal Intelligence Quotient | Descriptive | Observation | Similarities Vocabulary | 40 - 145 | - | Higher verbal abilities | Wechsler, 2018 |
| | Performance Intelligence Quotient | | | Block Design Visual Puzzles | 40 - 145 | - | Higher visual spatial abilities | |
| **Social Responsiveness Scale-Children (SRS-2)** | Symptom severity | Primary | Parent | 65 | 0 - 192 | 4 | Greater deficits in social responsiveness | Constantino & Gruber, 2012; Roeyers et al., 2015 |
| **Repetitive Behavior Scale-Revised (RBS-R)** | Repetitive behavior | Secondary | Parent | 43 | 0 - 129 | 4 | More severe repetitive behavior | Bodfish et al., 2000; Lam & Aman, 2007 |

| | | | | | | | | |
|---|---|---|---|---|---|---|---|---|
| **Screen for Child Anxiety Related Emotional Disorders (SCARED-NL)** | Anxiety | Secondary | Parent and self | 69 | 0 - 207 | 3 | Higher risk for anxiety disorders | Muris et al., 2007 |
| **Attachement Style Classification Questionnaire (ASCQ)**<br>**Anxious**<br>**Avoidant**<br>**Secure** | Attachment | Secondary | Self | 15 | Per subscale 5 - 25 | 5 | More anxious, avoidant or secure attachment toward their peers | Finzi et al., 2000 |
| **Attachement questionnaire**<br>**Anxious**<br>**Avoidant**<br>**Secure** | Attachment | Secondary | Self | 9 | Per subscale 3 - 21 | 7 | More anxiety, avoidance or trust toward their mother | Bosmans et al., 2014 |
