## Supplementary Table 6 for "Effects of multiple-dose intranasal oxytocin treatment on social responsiveness in children with autism: A randomized, placebo-controlled trial"

**Mean questionnaire scores for each treatment group and assessment session.**

For each questionnaire, raw mean scores and standard deviations are listed separately for each treatment group (oxytocin, placebo) and each assessment session (baseline, post, follow-up) of each phase (phase I and II).

| | Baseline | | | | Phase I (double-blind) | | | | | | | |
|---|---|---|---|---|---|---|---|---|---|---|---|---|
| | | | | | Post | | | | Follow-up | | | |
| | Oxytocin | | Placebo | | Oxytocin | | Placebo | | Oxytocin | | Placebo | |
| **Outcome Measure** | N | Mean ± SD | N | Mean ± SD | N | Mean ± SD | N | Mean ± SD | N | Mean ± SD | N | Mean ± SD |
| **Primary Outcome** | | | | | | | | | | | | |
| SRS informant based | 38 | 89.26 ± 21.66 | 39 | 87.87 ± 20.03 | 38 | 85.18 ± 23.56 | 38 | 83.66 ± 21.88 | 38 | 82.50 ± 25.58 | 39 | 82.49 ± 23.88 |
| **Secondary Outcomes - informant report** | | | | | | | | | | | | |
| RBS | 38 | 27.29 ± 15.24 | 39 | 26.64 ± 16.43 | 38 | 20.76 ± 13.27 | 38 | 19.50 ± 14.70 | 38 | 22.74 ± 14.19 | 39 | 22.23 ± 16.71 |
| SCARED Parent | 38 | 39.74 ± 21.74 | 39 | 45.15 ± 18.31 | 38 | 39.26 ± 21.51 | 38 | 40.16 ± 18.31 | 38 | 36.82 ± 21.08 | 39 | 39.77 ± 17.69 |
| **Secondary Outcomes - self report** | | | | | | | | | | | | |
| SCARED Child | 38 | 38.29 ± 20.99 | 39 | 39.05 ± 20.21 | 38 | 33.55 ± 20.67 | 39 | 35.67 ± 21.18 | 38 | 32.32 ± 20.57 | 39 | 32.69 ± 17.85 |
| ASCQ Secure | 38 | 19.97 ± 3.50 | 39 | 19.23 ± 2.78 | 38 | 19.08 ± 3.24 | 39 | 18.31 ± 3.42 | 38 | 18.61 ± 3.38 | 39 | 18.41 ± 3.75 |
| ASCQ Anxious | 38 | 13.45 ± 5.19 | 39 | 12.85 ± 4.15 | 38 | 12.53 ± 5.00 | 39 | 11.85 ± 3.80 | 38 | 12.45 ± 5.06 | 39 | 10.46 ± 3.42 |
| ASCQ Avoidant | 38 | 13.79 ± 4.00 | 39 | 14.08 ± 3.86 | 38 | 13.50 ± 3.91 | 39 | 13.00 ± 4.01 | 38 | 12.95 ± 4.38 | 39 | 12.62 ± 3.81 |
| Attachment Mother Anxiety | 38 | 4.74 ± 2.89 | 39 | 4.82 ± 2.78 | 38 | 5.50 ± 2.68 | 39 | 4.49 ± 2.02 | 38 | 5.58 ± 2.90 | 39 | 4.90 ± 2.33 |
| Attachment Mother Avoidance | 38 | 9.05 ± 4.76 | 39 | 7.97 ± 4.03 | 38 | 8.84 ± 5.01 | 39 | 7.15 ± 2.80 | 38 | 8.45 ± 4.72 | 39 | 7.31 ± 3.29 |
| Attachment Mother Secure | 38 | 16.76 ± 4.24 | 39 | 17.87 ± 3.13 | 38 | 17.00 ± 3.57 | 39 | 17.69 ± 3.37 | 38 | 17.42 ± 3.55 | 39 | 18.00 ± 2.93 |

| | Phase II (single-blind) | | | | | | | |
|---|---|---|---|---|---|---|---|---|
| | Post | | | | Follow-up | | | |
| | Oxytocin$_{first}$ | | Placebo$_{first}$ | | Oxytocin$_{first}$ | | Placebo$_{first}$ | |
| **Outcome Measure** | N | Mean ± SD | N | Mean ± SD | N | Mean ± SD | N | Mean ± SD |
| **Primary Outcome** | | | | | | | | |
| SRS informant based | 37 | 83.81 ± 25.14 | 38 | 77.37 ± 22.75 | 36 | 79.22 ± 24.28 | 36 | 78.14 ± 23.78 |
| **Secondary Outcomes - informant report** | | | | | | | | |
| RBS | 37 | 20.57 ± 16.51 | 38 | 19.13 ± 16.66 | 36 | 19.58 ± 15.55 | 36 | 17.08 ± 12.76 |
| SCARED Parent | 37 | 37.43 ± 22.57 | 38 | 37.92 ± 18.96 | 36 | 33.81 ± 21.88 | 36 | 35.75 ± 17.17 |
| **Secondary Outcomes - self report** | | | | | | | | |
| SCARED Child | 37 | 30.38 ± 20.47 | 39 | 31.82 ± 21.14 | 36 | 29.58 ± 20.36 | 37 | 29.19 ± 17.30 |
| ASCQ Secure | 37 | 18.32 ± 3.51 | 39 | 18.41 ± 3.85 | 36 | 18.19 ± 2.81 | 37 | 18.19 ± 3.41 |
| ASCQ Anxious | 37 | 12.24 ± 5.36 | 39 | 10.41 ± 3.75 | 36 | 11.83 ± 4.83 | 37 | 10.89 ± 4.21 |
| ASCQ Avoidant | 37 | 12.46 ± 3.58 | 39 | 12.77 ± 4.21 | 36 | 12.64 ± 3.94 | 37 | 13.24 ± 3.95 |
| Attachment Mother Anxiety | 37 | 5.70 ± 3.61 | 39 | 5.13 ± 3.29 | 36 | 5.72 ± 4.03 | 37 | 5.00 ± 2.99 |
| Attachment Mother Avoidance | 37 | 8.30 ± 5.06 | 39 | 6.85 ± 3.34 | 36 | 7.94 ± 5.23 | 37 | 7.16 ± 3.38 |
| Attachment Mother Secure | 37 | 16.97 ± 3.37 | 39 | 17.03 ± 4.20 | 36 | 17.14 ± 4.43 | 37 | 17.46 ± 3.12 |

SRS Social Responsiveness Scale, RBS-R Repetitive Behavior Scale-Revised, SCARED Screen for Child Anxiety Related Disorders, ASCQ Attachment Style Classification Questionnaire.
